# The impact of hospital waiting lists on labour market outcomes: quasi-experimental evidence from musculoskeletal treatment in England

**DOI:** 10.64898/2026.10.05.26364590

**Authors:** Hannah Aston, Vahé Nafilyan, Rachel Meacock, Daniel Ayoubkhani

## Abstract

We analyse the effect of waiting time for elective musculoskeletal treatment on labour market outcomes in England using hospital records for working-age adults who received treatment between April 2015 and March 2023 to Census, tax and benefit records. To address clinical prioritisation and unobserved severity, we leveraged provider-level waiting time as an instrument for individual waiting time and estimated two-stage least squares models adjusted for baseline labour market outcomes, demographic and socioeconomic characteristics, and provider, referral-time and procedure fixed effects. We find that longer waits reduce annual employee earnings, with effects persisting for up to five years. For an additional four weeks of waiting, the largest estimated annual losses range from £301 for osteoarthritis to £487 for other joint disorders. There was no evidence of effects on employment status or benefit receipt, suggesting that delays primarily operate through reduced productivity or hours worked among those who remain employed. Reducing waiting times may not only improve health outcomes but help patients maintain earnings and support economic growth.

## 1 Introduction

Musculoskeletal (MSK) conditions such as low back pain, neck pain, osteoarthritis and rheumatoid arthritis are a major source of disability and work-limiting ill health in high-income countries (1). They are highly prevalent, often chronic, and the leading contributor to disability worldwide, affecting physical functioning, mobility and the capacity to perform everyday activities, including work. In England, MSK conditions affect nearly one-third of the population and place substantial demands on the health system, accounting for a large share of primary care activity (2). Their economic importance arises not only from healthcare utilisation, but also from their potential to reduce labour supply through pain, functional limitation, sickness absence and reduced productivity.

A growing body of evidence links MSK conditions to poorer labour market outcomes. In England, individuals with MSK conditions have employment rates around 18 percentage points below those of the wider working-age population (3), consistent with sizeable reductions in labour force participation. Evidence from other high-income countries similarly shows that MSK conditions are associated with disability pension receipt, work limitation, job transitions and early exit from the labour market (4,5). These associations are suggestive of MSK conditions affecting labour supply through pain, reduced mobility, functional limitations, sickness absence and lower productivity while in work. The economic costs are likely to be largest among individuals with more severe conditions requiring hospital care: for England, the five-year earnings loss associated with people admitted to hospital for MSK conditions in 2023 alone has been estimated at £2.4 billion (6).

Timely treatment may mitigate these labour market losses by reducing pain, improving mobility and restoring functional capacity. Elective inpatient procedures for MSK conditions, particularly within trauma and orthopaedic services, have been associated with improvements in earnings and employment after treatment (6). However, access to elective treatment in England is increasingly delayed. The NHS constitutional standard that 92% of patients should start consultant-led treatment within 18 weeks has not been met since 2015/16, and waiting times have increased substantially since then, particularly following disruption during the COVID-19 pandemic (7). In addition to health losses experienced during the wait, the health returns to common elective surgeries such as hip and knee replacements have also been shown to significantly reduce as waiting times increase (8,9).

Despite growing evidence regarding the health consequences of extended waits (10–12), less is known about the implications for labour market outcomes, where impacts may occur both through health-related productivity losses whilst in employment and by weaking attachment to work. In Norway, extended waits for MSK treatments led to increased workplace absences and disability receipt (13). In England, a £375 million increase in earnings could be realised should waiting times for elective MSK inpatient care return to target levels (14). However, these studies have methodological limitations. Estimating the effects of longer waiting time is challenging because individual waiting times are not randomly assigned. Patients with more severe MSK conditions may be prioritised for earlier treatment, but severity is also likely to be associated with worse subsequent earnings, likelihood of employment and receipt of social security benefits. As a result, naïve comparisons of patients with shorter and longer waits may confound the effect of waiting time with differences in underlying health, which is difficult to measure well in administrative records.

This paper addresses that identification challenge by using an instrumental variable (IV) approach. We construct an IV based on provider-level waiting times to isolate variation in individual waiting times driven by local health-system congestion rather than patient severity or clinical prioritisation. We apply this approach to linked administrative data for over one million working-age adults who received elective inpatient MSK treatment in England between 2015 and 2023. We estimate the effect of waiting time on earnings, paid employment status and benefit receipt up to five years after treatment.

Our results show that longer waiting times led to economically meaningful reductions in post-treatment earnings that persist for several years. We do not find evidence of effects on employment status or benefit receipt, suggesting that delays primarily operate through reduced productivity or hours worked among those who remain employed.

This paper contributes to the literature in three ways. First, it provides new evidence on the labour market effects of treatment delays in a publicly funded, free-at-point-of-use healthcare system, complementing existing work focused on health outcomes. Second, it applies an IV strategy to address selection into waiting times, contributing to a small but growing literature using quasi-experimental variation to study healthcare rationing. Third, by leveraging rich administrative data with long follow-up, it quantifies the persistence and magnitude of earnings effects at scale.

## 2 Institutional context

The National Health Service (NHS) in England is a predominantly tax-funded healthcare system free at the point of use. Elective hospital services are delivered mainly by geographically dispersed public hospital organisations, known as NHS trusts, alongside some independent-sector providers treating NHS-funded patients. In this paper, a provider refers to the hospital organisation responsible for the admission rather than to an individual clinician. Patients generally enter consultant-led elective care following referral by a general practitioner or another authorised healthcare professional. For many elective services, patients have a right to choose among clinically appropriate providers, although realised provider choice may also reflect referral practices, geography, service availability and anticipated waiting times (15).

A referral starts a referral-to-treatment pathway, which may include an initial specialist consultation, diagnostic tests, imaging and non-surgical management before treatment is agreed. If the responsible specialist determines that inpatient or day-case treatment is required, a decision to admit is recorded and the patient enters the admitted waiting stage. Patients are prioritised according to clinical need, while the timing of admission also depends on hospital capacity, staffing, operating-theatre availability, bed availability and accumulated backlogs.

England applies a national access standard to consultant-led elective care. During the study period, the operational standard was that at least 92% of patients with an incomplete pathway should have waited no more than 18 weeks from referral to the start of treatment (16). This referral-to-treatment measure covers the entire pathway and should not be interpreted as the waiting-time measure used in this study. We measure only the interval from the date of decision-to-admit to the date of hospital admission because this component is recorded consistently in Hospital Episode Statistics Admitted Patient Care (HES APC) data for the study population.

Hospitals operate under common national access rules but face different local demand and capacity constraints. These constraints change over time with staffing, theatre schedules, available beds, cancellations and the size and composition of the existing waiting list. Consequently, otherwise similar patients may experience different waits depending on the provider and period in which they are treated. This provider-level variation is central to our identification strategy.

## 3 Data and empirical approach

### 3.1 Data Sources

We use linked administrative data for England to study the relationship between waiting times for elective inpatient MSK treatment and subsequent labour market outcomes. The analysis links HES APC records to the 2011 Census, death registrations, Pay As You Earn (PAYE) Real Time Information (RTI) records from His Majesty’s Revenue and Customs (HMRC), and the Benefits and Income dataset (BIDs) from the Department for Work and Pensions (DWP). Hospital records are used to identify elective inpatient admissions for MSK conditions, waiting times, treatment dates, providers and procedures. Census records provide baseline demographic and socioeconomic characteristics. HMRC PAYE records provide monthly information on employee status and earnings, covering employees only, while DWP records provide monthly information on benefit receipt.

HES is a national administrative database containing records of care delivered in NHS-funded care in England, including admitted patient care, outpatient appointments and historical accident and emergency attendances. The APC component records individual hospital episodes and includes information on diagnoses, procedures, admission and discharge dates, admission method, provider, residence and provider location (15). We use HES APC to identify elective inpatient admissions for MSK conditions, define the index treatment date, measure waiting time from decision-to-admit to admission, classify treatment providers, and control for procedure type. We used data from April 2015 to March 2023.

The 2011 Census provides a population-level snapshot of individuals and households in England and Wales in March 2011. Census data are collected every ten years and provide detailed demographic, socioeconomic, household and geographic information used for population statistics, policy planning, service allocation and research. We use the 2011 Census to measure pre-treatment individual characteristics, including sex, ethnicity, socioeconomic status, educational attainment, disability status, self-reported health and rural–urban residence. Although the 2021 Census has since been conducted, we use the 2011 Census as it is the most recently available at the start of our study period, thereby avoiding the inclusion of post-treatment socio-demographic information in our analysis.

Labour market outcomes are measured using HMRC PAYE and DWP benefit records. The PAYE system records gross earnings paid to employees for tax purposes in the UK. We accessed data that covered the period from April 2014 to March 2025. DWP records provide monthly information on benefit receipt from April 2014 to September 2021. Death registrations are used to censor follow-up after death.

The PAYE dataset was linked to the 2011 Census through the Office for National Statistics (ONS) Demographic Index (16), an anonymised, person-specific identifier compiled by ONS from various national administrative data sources with a 96.7% linkage rate. The HES and death registration datasets were joined to the 2011 Census by the NHS number, obtained via linkage of the 2011 Census and NHS Patient Registers 2011-2013 (17), with an 89.2% linkage rate. Records without an NHS number will not have linked and therefore will not be captured in this analysis.

### 3.2 Sample construction

The study population consists of adults aged 25–64 at the time of receiving NHS-funded elective inpatient treatment for a MSK condition in England between 1 April 2015 and 31 March 2023. MSK admissions are identified using International Classification of Diseases, 10th Revision (ICD-10) diagnosis codes and grouped into four clinically informed categories (see Supplementary Table 1 for code list): back, neck and MSK pain; osteoarthritis; other joint disorders; and other MSK conditions. Osteoporosis and immune-mediated inflammatory diseases are excluded because of low prevalence in the working-age inpatient population over the study period.

We restrict the sample to individuals who were enumerated in the 2011 Census (to obtain time-constant demographics), were usual residents of England at Census, could be linked to NHS and HMRC records, and had a recorded waiting time of no more than two years between the decision to admit and hospital admission. Longer recorded waits are excluded as likely data errors. For individuals with multiple qualifying admissions within a MSK condition group, we retain the first admission and define this as the index treatment. Therefore, each person can have only record per MSK condition group.

### 3.3 Outcomes and treatment variable

The treatment variable of interest is the individual waiting time for elective inpatient treatment, defined as the number of days between the decision to admit and the admission date. This captures the admitted-patient waiting period recorded in HES.

We study three labour market outcomes: annual gross employee pay, paid employment, and benefit receipt. Monthly earnings are measured using PAYE RTI records, calendarised to reflect the month in which work was done, deflated to 2023 prices using the Consumer Prices Index, and winsorised at the 99.9th percentile. Person-months without a PAYE record are assigned zero earnings. Annual gross employee pay is calculated as the sum of monthly PAYE earnings in each post-treatment year. Paid employment is defined as having positive PAYE earnings in the last month of the post-treatment year for that individual. Benefit receipt is defined as receiving any DWP benefit in the last month in the post-treatment year per individual; this outcome is observed until September 2021. Outcomes are measured one, two, three, four and five years after treatment, subject to data availability.

### 3.4 Identification strategy

A naïve comparison of patients with shorter and longer waiting times is unlikely to identify the causal effect of waiting. The main concern is that waiting time is endogenous. Patients with more severe MSK conditions may be prioritised for earlier treatment, while severity is also likely to predict subsequent earnings, employment and benefit receipt. Conversely, patients with less severe conditions may wait longer and have better labour market outcomes for reasons unrelated to waiting itself. A simple regression-based comparison of those with long waiting times to individuals with short waiting times may therefore confound the effect of waiting time with unobserved differences in underlying health, functional limitation and clinical prioritisation.

We address this problem using an IV approach. The instrument is a leave-one-out measure of average waiting time among patients treated by the same provider, in the same MSK condition group and in the same month of decision to admit, excluding the individual’s own waiting time. This instrument captures variation in local health-system congestion that affects individual waiting time but is plausibly unrelated to the individual patient’s unobserved severity or labour market trajectory, conditional on controls and fixed effects. Related studies have used similar provider- or area-level waiting-time instruments to isolate variation in waiting times that is less likely to reflect individual patient severity (13,18). The relevance condition requires that provider-level waiting times predict individual waiting times. This is plausible because patients treated by the same provider with a decision to admit in the same period are exposed to common constraints in capacity, staffing, theatre availability and backlog pressure. The leave-one-out construction avoids a mechanical relationship between the instrument and the individual’s own waiting time. The exclusion restriction requires that, conditional on observed characteristics, baseline labour market outcomes, provider fixed effects, calendar-time fixed effects and procedure fixed effects, provider-level congestion affects later labour market outcomes only through its effect on the individual’s own waiting time.

First, naïve ordinary least squares (OLS) models were estimated for individual-level waiting time across all condition groups separately (including all covariates and fixed effects described below). We then estimated two-stage least squares (2SLS) models separately for each MSK condition group, outcome and follow-up year.

In the first-stage equation, we modelled individual waiting time as a function of provider-level waiting times and a set of controls:

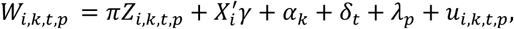

where *W_i_*_,*k*,*t*,*p*_is the waiting time for individual *i* treated by provider *k,* with a decision to admit in period *t* for procedure *p*, *Z_i_*_,*k*,*t*,*p*_is the leave-one-out provider-level waiting time average, *X_i_*is a vector of individual-level characteristics and baseline labour market outcomes, *α_k_*denotes provider fixed effects, *δ_t_*denotes calendar-time (year-month) fixed effects for the timing of the decision to admit, and *λ_p_*denotes procedure fixed effects based on three-digit Office of Population Censuses and Surveys Classification of Interventions and Procedures (OPCS) procedure codes (see supplementary table 2).

The second-stage equation is:

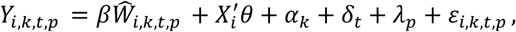

where *Y_i_*_,*k*,*t*,*p*_ is the labour market outcome for individual *i*, measured *k* years after treatment. The coefficient of interest is *β*, which gives the IV estimate of the effect of an additional day of waiting on the relevant labour market outcome. In the results, for ease of interpretation, we rescale estimates to show the effect of an additional four weeks of waiting. Patients without outcome or baseline data for the relevant year of modelling were excluded, as was anyone who died in that year.

### 3.5 Sensitivity Analyses

We conducted several sensitivity analyses to assess whether the main findings were robust to alternative modelling decisions and identifying assumptions. First, we compared the IV estimates with naïve OLS estimates to examine how accounting for the potential endogeneity of individual waiting time affected the results. We then estimated models including provider-by-procedure fixed effects to account more flexibly for differences in treatment patterns and capacity across providers, and models excluding individual-level characteristics to assess whether the estimates were sensitive to covariate adjustment. We also pooled all condition groups in a single model to examine whether the findings were robust to an alternative grouping strategy.

We additionally assessed the functional-form and monotonicity assumptions underlying the IV analysis. To assess whether the first-stage relationship was adequately represented by a linear model, we plotted predicted individual waiting times from linear, quadratic, cubic and natural cubic spline (with the spline fitted using three degrees of freedom and knots placed at the default quantiles of provider-level waiting time) specifications against provider-level waiting time and compared the shapes of the fitted relationships. This analysis examined whether meaningful non-linearity in the relationship could make the linear first-stage specification inappropriate. To assess monotonicity, we divided the instrument into ordered groups and plotted the empirical cumulative distribution function (CDF) of individual-level waiting time within each group. We examined whether higher values of the instrument consistently shifted the distribution towards longer individual waits, without substantial crossing of the CDF curves. This provided graphical evidence on whether provider-level waiting time affected individual waiting time in a consistent direction across patients.

All sensitivity analyses can be found in the supplementary materials.

All analyses were performed using R version 4.4.0. The 2SLS models were fitted using R’s ‘lfe’ package (19). Dataset preparation was carried out using Spark version 3.3.0.

## 4 Results

### 4.1 Sample characteristics and waiting times

The analytical sample comprised 1,019,897 working-age adults who received elective inpatient treatment for a MSK condition between 1 April 2015 and 31 March 2023 and satisfied the study inclusion criteria. A study sample flow diagram is shown in Supplementary Figure 1The mean age was 50.1 years at admission and 54.7% of patients were female. Median waiting time from decision-to-admit to admission was 69 days (interquartile range [IQR] 36–122). Individual-level waiting times varied across condition groups: patients with osteoarthritis had the longest median wait (83 days, IQR 44–147), followed by other joint disorders (69 days, IQR 38–120), other MSK conditions (65 days, IQR 35–114), and back, neck and MSK pain (58 days, IQR 31–108).

The median provider-level waiting time (the instrument) was 84 days (IQR 61-117) with longest median waits for patients with osteoarthritis (98 days IQR 71-141), followed by other joint disorders (84 days, IQR 63–115), other MSK conditions (80 days, IQR 59–109), and back, neck and MSK pain (75 days, IQR 54–104).

The most common three-digit OPCS procedure codes within each condition group are reported in Supplementary Table 2.

Table 1 compares observed characteristics for patients waiting less than 18 weeks from decision-to-admit to admission with those waiting 18 weeks or longer (see supplementary materials for Table 1 stratified by condition group). Differences were generally small but socio-economic status and MSK condition group had absolute standardised differences above 10%, conventionally defined as the threshold for indicating imbalance (20). Patients waiting at least 18 weeks were more likely to be in routine or semi-routine occupations, to have never worked or be long-term unemployed, and to have an osteoarthritis diagnosis.

**Table 1.** Distributions of covariates by time waited for treatment from decision-to-admit to admission (<18 weeks versus ≥ 18 weeks)

| Characteristic | Level | All (%) | < 18 weeks (%) | ≥ 18 weeks (%) | Absolute standardised difference (%) |
| --- | --- | --- | --- | --- | --- |
| Age | Mean age (years) | 50.06<br>(SD 10.46) | 49.90<br>(SD 10.39) | 50.56<br>(SD 10.68) | 6.22 |
| Sex | Females | 54.65 | 54.59 | 54.85 | 0.53 |
|  | Males | 45.35 | 45.41 | 45.15 |  |
| Ethnic group | Bangladeshi | 0.50 | 0.52 | 0.47 | 4.13 |
|  | Black African | 0.96 | 0.95 | 1.00 |  |
|  | Black Caribbean | 1.11 | 1.11 | 1.09 |  |
|  | Chinese | 0.20 | 0.20 | 0.18 |  |
|  | Indian | 2.15 | 2.19 | 2.02 |  |
|  | Mixed | 1.33 | 1.31 | 1.38 |  |
|  | Other | 1.91 | 1.93 | 1.84 |  |
|  | Pakistani | 1.40 | 1.42 | 1.32 |  |
|  | White British | 86.34 | 86.26 | 86.62 |  |
|  | White Other | 4.10 | 4.12 | 4.06 |  |
| Socio-economic status (at census) | Higher managerial, administrative and professional occupations | 8.63 | 8.83 | 7.99 | 10.02 |
|  | Intermediate occupations | 14.31 | 14.44 | 13.89 |  |
|  | Lower managerial, administrative and professional occupations | 22.47 | 22.77 | 21.48 |  |
|  | Lower supervisory and technical occupations | 8.10 | 8.11 | 8.08 |  |
|  | Never worked and long-term unemployed | 4.47 | 4.37 | 4.81 |  |
|  | No code required | 0.84 | 0.66 | 1.42 |  |
|  | Not classified | 3.35 | 3.17 | 3.94 |  |
|  | Routine occupations | 11.28 | 11.18 | 11.62 |  |
|  | Semi-routine occupations | 16.00 | 15.92 | 16.28 |  |
|  | Small employers and own account workers | 10.55 | 10.56 | 10.49 |  |
| Disability | Limited a little | 9.46 | 9.34 | 9.88 | 6.81 |
|  | Limited a lot | 6.41 | 6.25 | 6.93 |  |
|  | Not limited | 84.13 | 84.42 | 83.19 |  |
| Highest qualification | Apprenticeship | 3.63 | 3.63 | 3.63 | 9.00 |
|  | Level 1 qualification | 18.35 | 18.24 | 18.72 |  |
|  | Level 2 qualification | 18.62 | 18.59 | 18.74 |  |
|  | Level 3 qualification | 14.04 | 14.06 | 13.99 |  |
|  | Level 4 qualification or above | 26.13 | 26.51 | 24.90 |  |
|  | No code required | 0.84 | 0.66 | 1.42 |  |
|  | No qualifications | 13.90 | 13.82 | 14.14 |  |
|  | Other qualification | 4.49 | 4.50 | 4.46 |  |
| Rural / Urban residence | Rural | 18.09 | 17.84 | 18.89 | 2.72 |
|  | Urban | 81.91 | 82.16 | 81.11 |  |
| Self-reported health (at census) | Bad | 4.30 | 4.20 | 4.62 | 2.76 |
|  | Fair | 13.56 | 13.43 | 13.96 |  |
|  | Good | 42.05 | 42.14 | 41.76 |  |
|  | Very bad | 0.99 | 0.96 | 1.06 |  |
|  | Very good | 39.11 | 39.27 | 38.60 |  |
| MSK condition group | MSK Pain: Back, neck and MSK pain | 22.19 | 23.40 | 18.29 | 27.35 |
|  | MSK Pain: Osteoarthritis | 23.53 | 21.35 | 30.62 |  |
|  | MSK Pain: Other joint disorders | 30.28 | 30.46 | 29.70 |  |
|  | MSK Pain: Other MSK conditions | 24.00 | 24.80 | 21.39 |  |
| Index of Multiple Deprivation (IMD) Quintile Group | 1 (most deprived) | 19.13 | 19.02 | 19.47 | 4.16 |
|  | 2 | 19.92 | 19.80 | 20.28 |  |
|  | 3 | 20.59 | 20.46 | 20.99 |  |
|  | 4 | 20.68 | 20.75 | 20.44 |  |
|  | 5 (least deprived) | 19.41 | 19.70 | 18.48 |  |

**Table 2.** First-stage relationship between provider-level waiting time and individual waiting time.

| MSK condition group | Outcome | Coefficient on provider-level waiting time | Standard error | Mean individual waiting time (days) | Observations (n) | First-stage F-statistic |
| --- | --- | --- | --- | --- | --- | --- |
| Back, neck and MSK pain | Gross pay | 0.955 | 0.007 | 85 | 217,690 | 16,265 |
|  | Employment | 0.981 | 0.004 | 86 | 218,476 | 64,066 |
|  | Benefit receipt | 0.851 | 0.060 | 81 | 48,292 | 204 |
| Osteoarthritis | Gross pay | 0.950 | 0.009 | 113 | 228,359 | 12,444 |
|  | Employment | 0.981 | 0.004 | 114 | 219,663 | 52,042 |
|  | Benefit receipt | 0.741 | 0.085 | 97 | 37,627 | 76 |
| Other MSK conditions | Gross pay | 0.962 | 0.007 | 89 | 236,369 | 18,319 |
|  | Employment | 0.988 | 0.006 | 90 | 235,292 | 31,613 |
|  | Benefit receipt | 0.774 | 0.059 | 81 | 35,882 | 170 |
| Other joint disorders | Gross pay | 0.960 | 0.008 | 95 | 297,909 | 13,833 |
|  | Employment | 0.990 | 0.004 | 97 | 300,145 | 74,999 |
|  | Benefit receipt | 0.779 | 0.063 | 85 | 36,461 | 155 |

**Notes:** The dependent variable is individual waiting time from decision-to-admit to admission. The excluded instrument is the leave-one-out mean waiting time for patients treated by the same provider, in the same MSK condition group, and in the same calendar year and month of decision to admit. All specifications include the full set of covariates, baseline labour market outcomes, provider fixed effects, calendar-time fixed effects and three-digit OPCS procedure fixed effects used in the second-stage models. Standard errors are clustered by provider and year-month of decision to admit. The first-stage F-statistic tests the excluded instrument. For first-stage output and F-statistics for years two to five, see supplementary materials. Coefficients and standard errors are expressed on a 'per day' basis.

### 4.2 First-stage regression results

The first-stage model indicates that the leave-one-out provider-level waiting-time measure is a strong predictor of individual waiting time. The first-stage F-statistics for the excluded instrument are strong for all outcomes, considerably above the widely accepted threshold of 50 due to the near-population level sample (21).

### 4.3 2SLS estimates of the effects of waiting times on earnings

Figure 1 reports the 2SLS estimates of the effect of an additional four weeks of waiting on annual gross employee pay in each of the five post-treatment years. Across all four MSK condition groups, longer waiting times are associated with lower subsequent earnings. The pattern is economically meaningful and persistent: estimated losses are already apparent in the first post-treatment year, generally increase over the first three to four years, and remain significant in year five.

**Figure 1.**
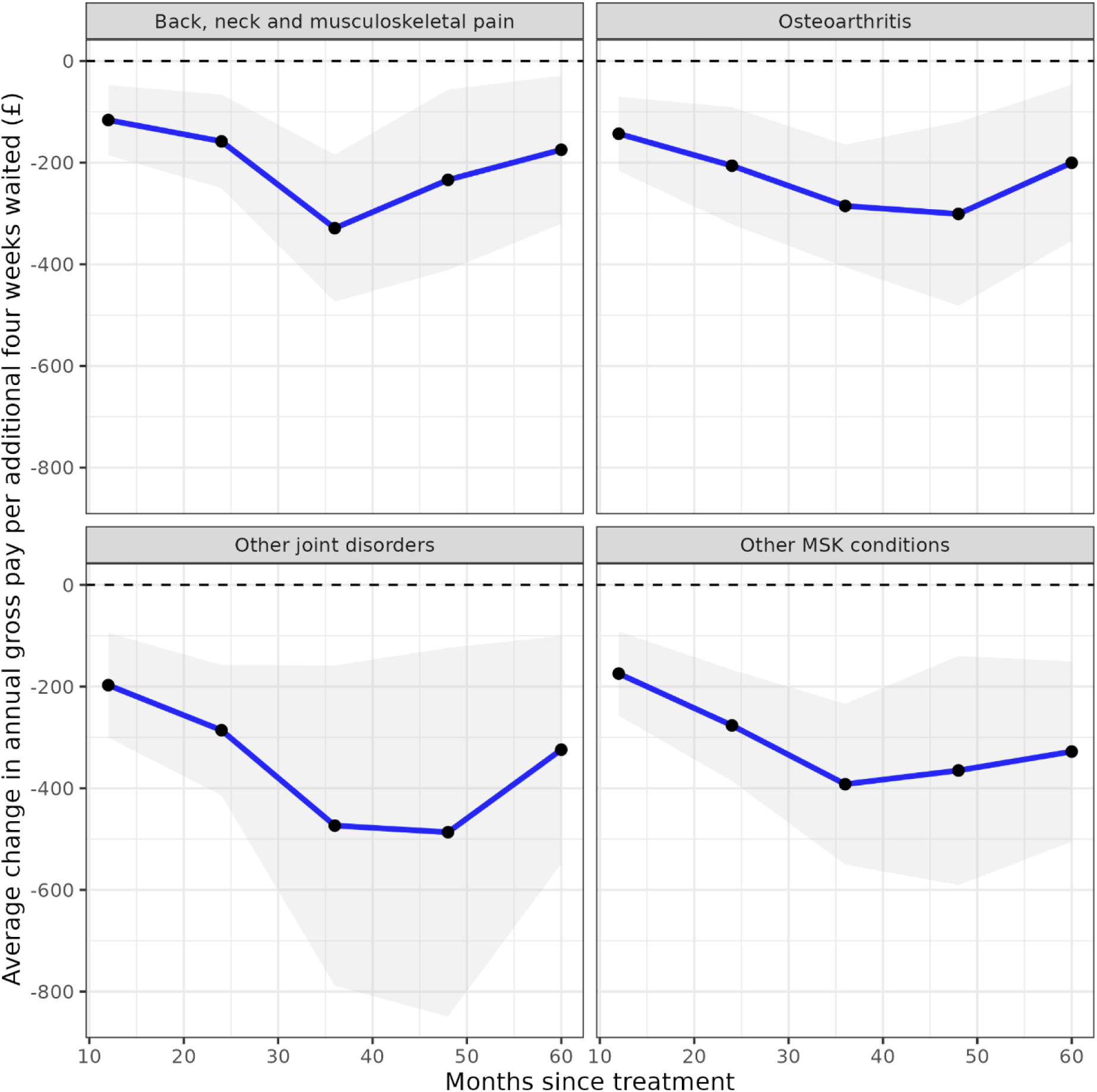
Estimated relationship between waiting time and gross employee pay up to five years after treatment, stratified by MSK condition group *Notes: Estimates are adjusted for age at index date, sex, ethnic group,* socio-economic status, disability, highest qualification, rural or urban residence, self-reported general health and IMD quintile group, baseline pay, and fixed effects (provider, referral year and month, and three digit OPCS-4 code). *Shaded areas are 95% confidence intervals*.

The largest estimated earnings effects are observed for patients with other joint disorders. For this group, an additional four weeks of waiting is associated with £197 lower annual pay in the first year after treatment (95% confidence interval [CI]: £94 to £300), rising to £487 in year four (95% CI: £124 to £849). The estimated loss remains sizeable in year five, at £324 (95% CI: £100 to £548).

For osteoarthritis, the estimated loss is £143 in year one (95% CI: £70 to £216), increasing to £301 by year four (95% CI: £120 to £481), before declining to £200 in year five (95% CI: £47 to £185). For back, neck and MSK pain, the estimated loss is £116 in year one (95% CI: £47 to £185), peaks at £329 in year three (95% CI: £185 to £473), and is £175 in year five (95% CI: £29 to £320).

For other MSK conditions, the estimated loss is £174 in year one (95% CI: £92 to £257), increases to £392 in year three (95% CI: £234 to £550), and remains at £328 in year five (95% CI: £151 to £505). Taken together, these estimates suggest that treatment delays are associated with reductions in earnings that persist well beyond the treatment episode, rather than short-lived earnings disruption around the admission itself.

### 4.4 Heterogeneity in earnings effects

Supplementary Figures 2–4 examine heterogeneity in the estimated earnings effects by sex, age group and area deprivation (Index of Multiple Deprivation [IMD] quintile group). The point estimates are generally more negative for younger adults than for older adults, for males than females–particularly among those with other joint disorders and other MSK conditions—and for patients living in less deprived areas relative to those in more deprived areas. he largest differences between males and females were observed for other joint disorders. When expressed as a percentage of baseline annual earnings, the estimated reductions were 18% for females and 27% for males. Differences by age were also most pronounced for other joint disorders, with estimated reductions equivalent to 13% of baseline annual earnings among adults aged 55 years and over and 24% among those aged 35 to 44 years. These patterns are consistent with larger potential earnings losses among groups with stronger pre-treatment labour market attachment or higher baseline earnings. However, 95% CIs are wide and overlap substantially across most subgroup comparisons, so the heterogeneity results should be interpreted as descriptive evidence on numerical patterns of estimates rather than inferential evidence of differential treatment effects.

### 4.5 Employment and benefit receipt

In contrast to the earnings results, there is no statistical evidence that longer waiting times affect paid employment status or benefit receipt over the five years after treatment. For both outcomes, the 95% CIs include zero at each follow-up year and across all MSK condition groups (Figures 2 and 3). This pattern suggests that the estimated labour market consequences of longer waits operate primarily through reduced earnings among those with PAYE employment, rather than through measurable changes in the probability of being in paid employment or receiving benefits.

**Figure 2.**
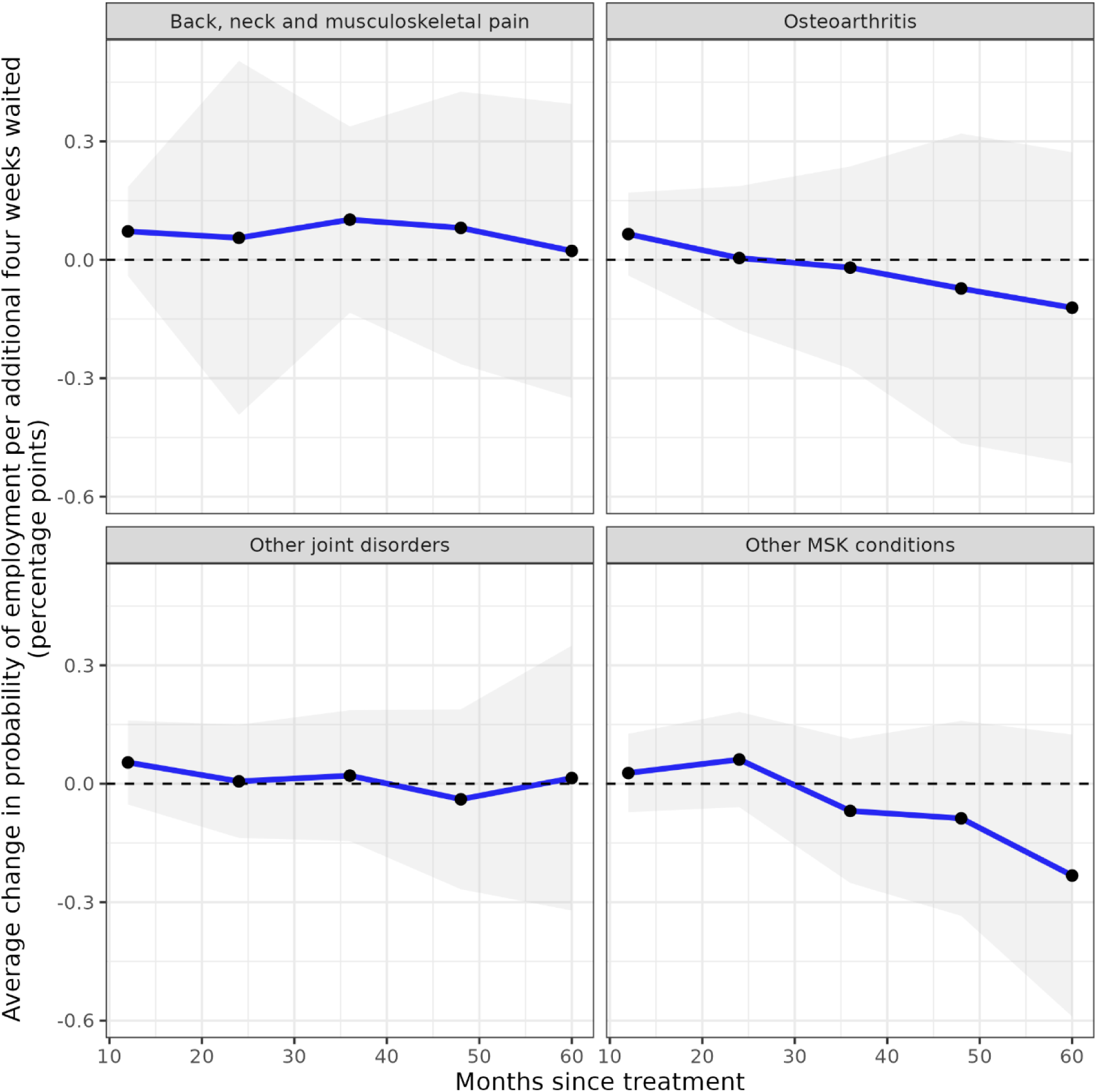
Estimated relationship between waiting time and employment status up to five years after treatment, stratified by MSK condition group *Notes: Estimates are adjusted for age at index date, sex, ethnic group,* socio-economic status, disability, highest qualification, rural or urban residence, self-reported general health and IMD quintile group, baseline employee status, and fixed effects (provider, referral year and month, and three digit OPCS-4 code). *Shaded areas are 95% confidence intervals*.

**Figure 3.**
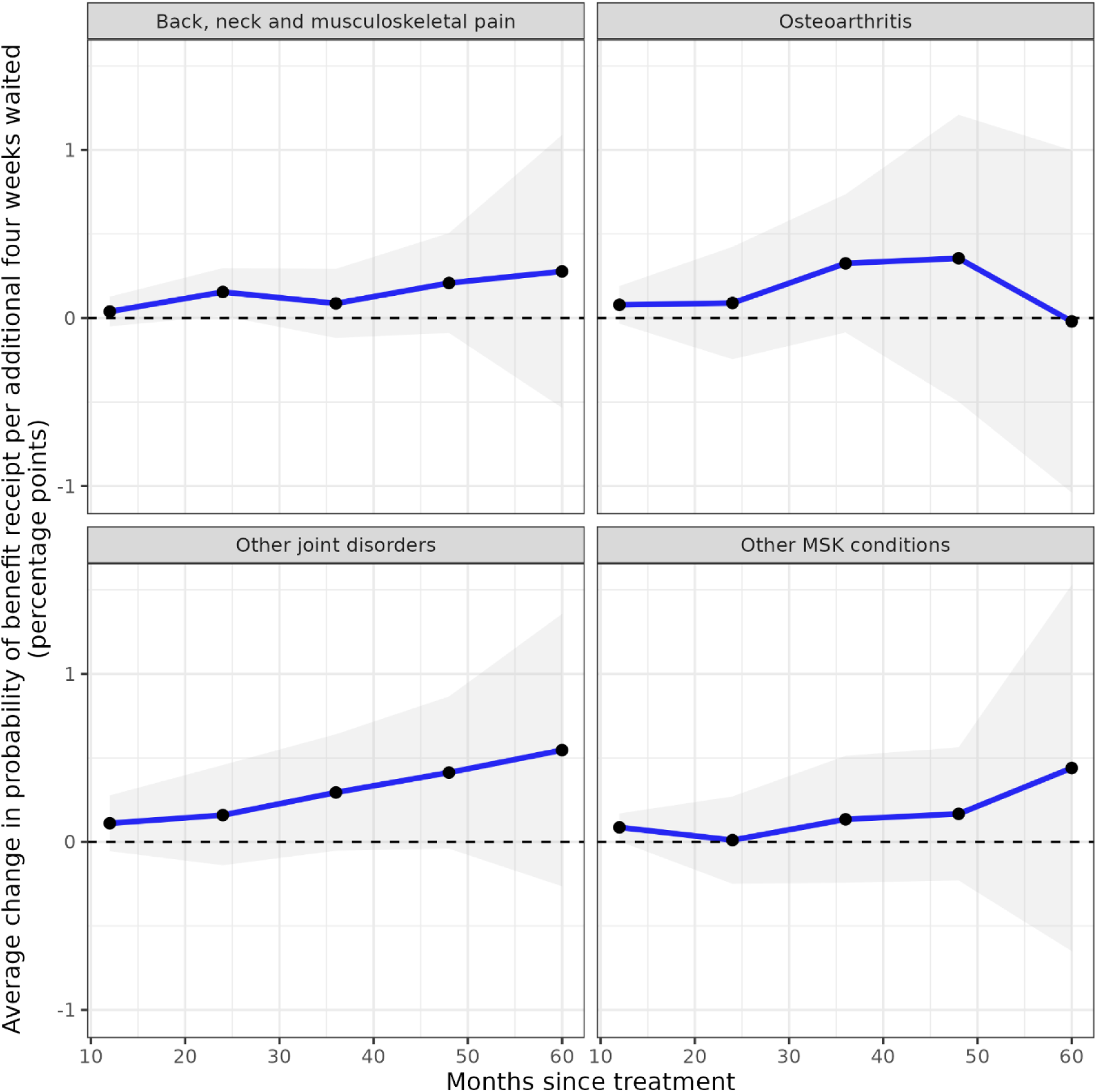
Estimated relationship between waiting time and benefit receipt up to five years after treatment, stratified by MSK condition group *Notes: Estimates are adjusted for age at index date, sex, ethnic group,* socio-economic status, disability, highest qualification, rural or urban residence, self-reported general health and IMD quintile group, baseline benefit receipt status, and fixed effects (provider, referral year and month, and three digit OPCS-4 code). *Shaded areas are 95% confidence intervals*.

### 4.6 Sensitivity analyses

The main findings were robust to alternative model specifications, including the use of provider-by-procedure fixed effects, the exclusion of individual-level covariates, and the estimation of a combined model across condition groups. The direction and magnitude of the estimated effects were broadly consistent with the primary analysis (Supplementary Appendix A, Figures A1–A16).

## 5 Discussion

### 5.1 Principal findings

We found that extended waiting times for elective inpatient MSK treatment were associated with lower pay up to five years post-treatment. The largest decreases were observed among patients treated for other joint disorders, who experienced reductions of more than £1,760 per additional four weeks waited over the five-year period after treatment; the most common procedure in this group was therapeutic endoscopic operations on semilunar cartilage. Substantial declines were also observed among patients with osteoarthritis, back and neck pain, and other MSK conditions. These estimates imply that even moderate increases in waiting time can generate long-lasting earnings losses. The persistence of these effects suggests that treatment delays may have consequences beyond short-term disruption around admission.

These findings are important given the high prevalence of MSK conditions and their substantial effect on working capacity and labour market participation. Although the estimated losses are relatively modest at an individual level, they could translate into sizeable reductions in aggregate earnings, productivity and tax receipts across the large number of working-age patients receiving MSK treatment.

### 5.2 Mechanisms

Whilst we found that increased waiting time led to a reduction in annual earnings, there was no statistical evidence of a relationship between waiting time and either employment or benefit receipt. Taken together, these findings suggest that extended waiting times primarily affect labour market engagement through reduced earnings among those who remain employed, rather than through changes in employment status or movement onto benefits. Possible mechanisms include reduced working hours, sickness absence, lower productivity, or movement into less demanding and lower-paid roles, although these could not be distinguished using our data.

Several mechanisms could account for persistent earnings effects. Longer waits may prolong pain and functional limitation before treatment, increase sickness absence, weaken attachment to an existing job or induce movement into less physically demanding and lower-paid work. Delay may also reduce the health gain from treatment or make recovery less complete, allowing effects generated during the waiting period to continue after admission (8,9). The available data do not distinguish between these channels. The pattern is nevertheless consistent with dynamic labour market effects in which a temporary period of reduced work capacity has consequences beyond the treatment episode. The findings may generalise most readily to other painful or mobility-limiting conditions where delays restrict a person’s capacity to work without necessarily causing them to leave employment. Generalisability to other conditions and healthcare systems is less certain because treatment pathways, employment protections and waiting-list arrangements differ.

### 5.3 Comparison with prior evidence

Our findings complement evidence that extended waits for elective care worsen patients’ physical and mental health (8,10–12,22), increase risk of mortality in both elective (23,24) and emergency settings (25), and affect patient outcomes (26,27). Evidence also suggests that longer waits can influence labour market participation, including impacts on earnings and work capacity, though findings vary across conditions and settings (13,14,28,29). The closest economic comparison is a Norwegian study of orthopaedic surgery, which used variation in queue congestion and found persistent increases in work absence and permanent disability receipt, particularly among people already on sick leave at referral (13). Our finding of lower earnings but no detectable effect on employment or benefit receipt differs in the margin of labour supply affected. This may reflect differences between England and Norway in sickness-insurance coverage, employment protection, disability-benefit eligibility and waiting-list management, as well as differences in study populations and outcome definitions.

This study extends the literature by estimating labour market effects for more than one million working-age patients treated in a tax-funded health system and by following earnings for up to five years post-treatment. To our knowledge, this is the first study to examine the relationship between waiting times and labour market outcomes for MSK patients in England using individual-level administrative data and an IV approach. Other English studies have attempted to estimate the economic benefits of reducing elective waits, but these analyses relied on aggregated data (30) or were unable to fully capture heterogeneity in treatment effects according to time waited for treatment due to only adjusting for age (14). The individual-level linkage used here permits adjustment for baseline labour market outcomes and observed demographic and socioeconomic characteristics, while the IV design addresses prioritisation on unobserved patient severity under the identifying assumptions described below.

### 5.4 Strengths and limitations

A key strength is the use of novel linked administrative data comprising records for over one million patients with an MSK diagnosis. By linking national records on healthcare use, earnings, employment and benefit receipt, we could examine long-term outcomes at multiple timepoints at an individual level without relying on self-reported information or survey samples, reducing the risk of recall error and survey non-response bias. To our knowledge, this is one of the largest studies internationally to quantify the labour market consequences of receiving elective inpatient treatment for an MSK condition; previous studies have been limited to aggregated data or smaller cohorts (13,31). A further strength is the use of a strong provider-level waiting time instrument, supported by large first-stage F-statistics. Graphical checks also supported a linear first-stage relationship and the monotonicity assumption. Lastly, our system-wide instrument was a strong predictor of individual waits and is unlikely to be related to individual labour market status, allowing us to account for individual condition severity and subsequent omitted variable bias in our modelling and produce causal estimates.

Our study also has limitations. The HES dataset contains only records for patients who received treatment during the study period. Therefore, individuals still awaiting treatment at the end of the study period are not included, and the observed cohort sizes will be smaller than the actual number of patients waiting for treatment during the study period. This may result in bias if there is time-dependent heterogeneity in the relationship between waiting time and labour market outcomes, particularly in the more recent past.

As the HES APC dataset does not consistently record the time waited from referral to treatment, our analysis focuses on waiting time from decision-to-admit to admission. Earlier stages of the pathway may include referral, specialist assessment and diagnostic testing, and not every referral results in a decision to admit for inpatient treatment. We therefore examine the period after clinicians have determined that inpatient treatment is required, up to the point at which treatment begins. Although this is a clinically relevant and consistently recorded part of the pathway, we cannot assess delays before the decision to admit. This may introduce bias if the time from referral to decision-to-admit is related to labour market outcomes.

The PAYE RTI dataset captures only employees. Although self-employed individuals (who make up approximately 13% of the UK workforce (32)) are included in the study population, they appear in the data as receiving £0 pay and are therefore classified as not being paid employees when deriving the pay and employment outcome variables for this analysis. Similarly, individuals who are employed but not currently receiving pay via PAYE, for example, those on maternity leave or long-term sickness absence, are also categorised as not being paid employees. In addition, we did not have reliable information on hours worked or those receiving sick pay. Consequently, we were unable to infer the extent to which the estimated relationship between waiting time and gross employee pay is attributable to changes in working hours versus other factors (e.g. changes in hourly wages).

Despite the overall cohort size, the study population represents a narrow and selective subgroup of the wider inpatient population, as it includes only individuals who received elective inpatient treatment for MSK conditions. This focus means the cohort is inherently restricted to people with specific diagnoses and sufficient clinical severity to warrant inpatient care. As a result, the findings may not generalise to elective inpatients being treated for conditions other than MSK, or MSK patients treated in outpatient or community settings (likely to be those with less severe illness).

### 5.5 External validity and implications

The estimates apply most directly to working-age adults who eventually received elective inpatient treatment for MSK conditions in England and whose waits were responsive to provider congestion. They may be informative for other painful or mobility-limiting conditions, but generalisability to outpatient care, untreated patients and countries with different financing, sickness-insurance and employment-protection arrangements is uncertain. Subject to the IV assumptions, the findings indicate that the opportunity-cost of elective queues extends beyond health losses to patients’ subsequent earnings. Decisions about expanding capacity should therefore consider potential labour market benefits alongside health gains and the resource costs of reducing waits, rather than treating the estimates as evidence that any particular waiting-time target is optimal.

## 6 Conclusions

This study provides evidence on the labour market consequences of extended waiting times for elective inpatient MSK treatment in England, using a national, linked administrative dataset and an IV approach to guard against omitted variable bias. We find that longer waits are consistently associated with substantial and persistent reductions in earnings in the five years following treatment, but no evidence of a relationship between waiting time and employment status or benefit receipt. These results suggest that prolonged waits may primarily limit individuals’ capacity to maintain earnings (for example, due to working fewer hours or moving into less demanding and lower paid roles) rather than pushing them out of employment entirely. Our study highlights the wider economic consequences of rising elective care backlogs in the UK and reinforce the importance of reducing waiting times, not only for improving health outcomes, but also for supporting long-term labour market participation and ultimately economic growth. Further work is needed to understand the causal mechanisms underpinning our statistical findings, and to investigate the relationship between waiting times and labour market outcomes for MSK patients treated outside of hospital inpatient settings.

## Availability of data and materials

The source data used in this study is subject to controlled access due to its sensitive nature and legal constraints.

## Authors’ contributions

HA, DA and VN conceptualised and designed the study. HA, DA prepared the study data and performed the statistical analysis. All authors contributed to interpretation of the results. HA and DA were responsible for the first draft of the manuscript. All authors contributed to critical revision of the manuscript. All authors approved the final manuscript.

## Ethics statement

Ethical approval was obtained from the National Statistician’s Data Ethics Advisory Committee (NSDEC23(18)).

