## Supplementary Materials for "The impact of hospital waiting lists on labour market outcomes: quasi-experimental evidence from musculoskeletal treatment in England"

**Supplementary Table 1. MSK ICD-10 code list**

| **MSK condition group** | **ICD-10 Code** |
| --- | --- |
| Back, neck and musculoskeletal pain | M954, M964, M923, M548, M943, M519, M860, M902, M921, M955, M942, M920, M501, M965, M904, M471, M531, M410, M960, M481, M494, M873, M436, M871, M913, M479, M889, M906, M864, M931, M508, M485, M926, M900, M503, M429, M498, M472, M905, M966, M941, M930, M868, M514, M421, M901, M414, M922, M509, M895, M908, M435, M951, M961, M430, M404, M478, M495, M925, M530, M546, M968, M538, M911, M438, M402, M413, M903, M907, M401, M929, M484, M545, M412, M863, M480, M932, M893, M544, M948, M541, M433, M439, M919, M962, M470, M928, M870, M861, M869, M892, M483, M431, M872, M403, M419, M434, M927, M938, M963, M952, M940, M866, M865, M533, M493, M924, M502, M878, M532, M899, M880, M405, M539, M518, M894, M912, M910, M415, M896, M540, M543, M890, M549, M400, M939, M482, M949, M542, M432, M958, M969, M898, M489, M953, M888, M891, M879, M420, M411, M512, M918, M511, M513, M500, M862, M418, M510, M950, M959 |
| Osteoarthritis | M154, M183, M162, M172, M173, M159, M152, M180, M175, M169, M198, M189, M163, M153, M199, M171, M150, M170, M161, M185, M179, M158, M181, M165, M174, M190, M167, M160, M182, M166, M192, M164, M151, M184, M191 |
| Other joint disorders | M231, M249, M254, M224, M244, M251, M210, M200, M240, M214, M203, M257, M215, M204, M239, M250, M255, M256, M232, M247, M243, M206, M253, M234, M205, M220, M236, M201, M228, M246, M219, M223, M233, M221, M202, M212, M242, M259, M222, M258, M218, M229, M217, M235, M213, M241, M211, M252, M230, M216, M248, M245, M238 |
| Other MSK conditions | G551, M730, M652, M753, M611, M759, M670, M602, M779, M600, M761, M714, M990, M650, M751, M678, M726, M654, M992, M672, M796, M752, M999, M613, M763, M619, M997, M708, M361, M773, M630, M700, M359, M673, M998, M719, M364, M664, M750, M758, M762, M615, M731, M614, M713, M772, M718, M710, M755, M628, M996, M612, M709, M768, M764, M794, M991, M797, M629, M778, M701, M760, M712, M707, M721, M769, M679, M651, M995, M711, M799, M771, M770, M722, M610, M674, M680, M703, M765, M671, M795, M625, M653, M791, M633, M663, M638, M775, M626, M738, M702, M624, M793, M623, M728, M798, M608, M363, M661, M754, M756, M659, M792, M658, M631, M622, M767, M621, M790, M357, M706, M724, M665, M993, M766, M660, M704, M729, M632, M601, M620, M774, M662, M705, M688, M994, M720, M715 |

**Supplementary Table 2. Top 20 most common three-digit OPCS-4 codes by MSK condition group**

| **MSK condition group** | **Three-digit OPCS-4 Code** | **Number of patients** | **Percentage (%)** |
| --- | --- | --- | --- |
| Back, neck and musculoskeletal pain | A57 | 48,922 | 21.7 |
|  | A52 | 40,315 | 17.9 |
|  | V54 | 36,003 | 16.0 |
|  | V25 | 16,452 | 7.3 |
|  | A73 | 14,606 | 6.5 |
|  | W90 | 10,286 | 4.6 |
|  | V22 | 7,957 | 3.5 |
|  | V33 | 6,557 | 2.9 |
|  | X37 | 4,535 | 2.0 |
|  | V29 | 3,957 | 1.8 |
|  | W08 | 3,503 | 1.6 |
|  | V48 | 2,555 | 1.1 |
|  | V67 | 1,890 | 0.8 |
|  | V38 | 1,593 | 0.7 |
|  | W84 | 1,119 | 0.5 |
|  | X29 | 1,116 | 0.5 |
|  | W83 | 1,070 | 0.5 |
|  | V40 | 938 | 0.4 |
|  | W38 | 859 | 0.4 |
|  | X11 | 829 | 0.4 |
| Osteoarthritis | W40 | 50,728 | 21.2 |
|  | W90 | 48,931 | 20.4 |
|  | W38 | 37,465 | 15.6 |
|  | W94 | 19,500 | 8.1 |
|  | W37 | 13,214 | 5.5 |
|  | W58 | 12,458 | 5.2 |
|  | W06 | 5,134 | 2.1 |
|  | W83 | 4,589 | 1.9 |
|  | W62 | 3,702 | 1.5 |
|  | W59 | 3,083 | 1.3 |
|  | W89 | 3,009 | 1.3 |
|  | W82 | 2,783 | 1.2 |
|  | W80 | 2,519 | 1.1 |
|  | O29 | 2,486 | 1.0 |
|  | W93 | 1,969 | 0.8 |
|  | W08 | 1,813 | 0.8 |
|  | W41 | 1,788 | 0.7 |
|  | W87 | 1,581 | 0.7 |
|  | W84 | 1,528 | 0.6 |
| Other joint disorders | W82 | 117,356 | 38.1 |
|  | W90 | 33,882 | 11.0 |
|  | W74 | 23,759 | 7.7 |
|  | W15 | 11,671 | 3.8 |
|  | W79 | 11,436 | 3.7 |
|  | W13 | 10,314 | 3.3 |
|  | W59 | 9,564 | 3.1 |
|  | O27 | 7,744 | 2.5 |
|  | W83 | 7,394 | 2.4 |
|  | W84 | 5,293 | 1.7 |
|  | W85 | 4,569 | 1.5 |
|  | W77 | 4,340 | 1.4 |
|  | W08 | 4,047 | 1.3 |
|  | W71 | 4,016 | 1.3 |
|  | W03 | 3,600 | 1.2 |
|  | W80 | 3,416 | 1.1 |
|  | W87 | 3,321 | 1.1 |
|  | W89 | 3,140 | 1.0 |
|  | W78 | 3,047 | 1.0 |
|  | A73 | 2,172 | 0.7 |
| Other MSK conditions | O29 | 42,691 | 17.5 |
|  | T59 | 22,637 | 9.3 |
|  | T72 | 19,729 | 8.1 |
|  | T52 | 19,565 | 8.0 |
|  | T79 | 16,601 | 6.8 |
|  | W90 | 12,605 | 5.2 |
|  | T62 | 11,091 | 4.5 |
|  | T74 | 9,043 | 3.7 |
|  | W78 | 8,719 | 3.6 |
|  | A57 | 7,214 | 3.0 |
|  | T69 | 5,561 | 2.3 |
|  | X29 | 3,786 | 1.6 |
|  | W84 | 3,291 | 1.3 |
|  | A73 | 3,054 | 1.3 |
|  | T57 | 2,979 | 1.2 |
|  | W08 | 2,784 | 1.1 |
|  | X37 | 2,734 | 1.1 |
|  | W91 | 2,708 | 1.1 |
|  | W69 | 2,606 | 1.1 |
|  | T54 | 2,558 | 1.0 |

**Supplementary Table 3. Distributions of covariates by time waited for treatment (<18 weeks versus ≥ 18 weeks) for Back, neck and musculoskeletal pain**

| **Characteristic** | **Level** | **All (%)** | **< 18 weeks (%)** | **≥ 18 weeks (%)** | **Absolute standardised difference (%)** |
| --- | --- | --- | --- | --- | --- |
| Age | Mean age (years) | 48.30  (SD 10.23) | 48.20  (SD 10.22) | 48.73 (SD 10.26) | 5.19 |
| Sex | Females | 58.05 | 57.64 | 59.74 | 4.27 |
|  | Males | 41.95 | 42.36 | 40.26 |  |
| Ethnic group | Bangladeshi | 0.72 | 0.70 | 0.80 | 10.76 |
|  | Black African | 1.20 | 1.17 | 1.31 |  |
|  | Black Caribbean | 1.04 | 1.05 | 0.99 |  |
|  | Chinese | 0.19 | 0.19 | 0.18 |  |
|  | Indian | 1.96 | 1.94 | 2.07 |  |
|  | Mixed | 1.46 | 1.44 | 1.57 |  |
|  | Other | 2.27 | 2.21 | 2.54 |  |
|  | Pakistani | 1.85 | 1.80 | 2.04 |  |
|  | White British | 84.77 | 84.99 | 83.87 |  |
|  | White Other | 4.54 | 4.52 | 4.64 |  |
| Socio-economic status | Higher managerial, administrative and professional occupations | 7.26 | 7.46 | 6.44 | 11.39 |
|  | Intermediate occupations | 14.25 | 14.38 | 13.69 |  |
|  | Lower managerial, administrative and professional occupations | 20.83 | 21.23 | 19.18 |  |
|  | Lower supervisory and technical occupations | 7.88 | 7.90 | 7.79 |  |
|  | Never worked and long-term unemployed | 6.19 | 5.93 | 7.30 |  |
|  | No code required | 0.71 | 0.61 | 1.15 |  |
|  | Not classified | 3.38 | 3.27 | 3.85 |  |
|  | Routine occupations | 12.32 | 12.19 | 12.86 |  |
|  | Semi-routine occupations | 17.27 | 17.08 | 18.04 |  |
|  | Small employers and own account workers | 9.91 | 9.96 | 9.71 |  |
| Disability | Limited a little | 11.16 | 10.94 | 12.06 | 14.09 |
|  | Limited a lot | 8.95 | 8.64 | 10.24 |  |
|  | Not limited | 79.89 | 80.42 | 77.70 |  |
| Highest qualification | Apprenticeship | 2.99 | 3.03 | 2.79 | 9.79 |
|  | Level 1 qualification | 19.05 | 18.97 | 19.38 |  |
|  | Level 2 qualification | 19.18 | 19.22 | 19.01 |  |
|  | Level 3 qualification | 13.95 | 14.04 | 13.57 |  |
|  | Level 4 qualification or above | 23.16 | 23.54 | 21.55 |  |
|  | No code required | 0.71 | 0.61 | 1.15 |  |
|  | No qualifications | 16.02 | 15.71 | 17.33 |  |
|  | Other qualification | 4.95 | 4.88 | 5.22 |  |
| Rural / Urban residence | Rural | 16.56 | 16.62 | 16.28 | 0.93 |
|  | Urban | 83.44 | 83.38 | 83.72 |  |
| Self-reported health | Bad | 6.13 | 5.91 | 7.07 | 7.94 |
|  | Fair | 15.98 | 15.74 | 16.95 |  |
|  | Good | 41.93 | 42.02 | 41.56 |  |
|  | Very bad | 1.53 | 1.45 | 1.86 |  |
|  | Very good | 34.43 | 34.88 | 32.57 |  |
| Index of Multiple Deprivation (IMD) Quintile Group | 1 (most deprived) | 22.08 | 21.60 | 24.06 | 5.79 |
|  | 2 | 21.26 | 21.10 | 21.89 |  |
|  | 3 | 20.04 | 20.19 | 19.42 |  |
|  | 4 | 18.91 | 19.09 | 18.16 |  |
|  | 5 (least deprived) | 17.44 | 17.75 | 16.16 |  |

**Supplementary Table 4. Distributions of covariates by time waited for treatment from decision-to-admit to admission (<18 weeks versus ≥ 18 weeks) for Osteoarthritis**

| **Characteristic** | **Level** | **All (%)** | **< 18 weeks (%)** | **≥ 18 weeks (%)** | **Absolute standardised difference (%)** |
| --- | --- | --- | --- | --- | --- |
| Age | Mean age (years) | 55.84  (SD 7.61) | 55.58  (SD 7.74) | 56.43 (SD 7.27) | 11.33 |
| Sex | Females | 56.88 | 57.08 | 56.44 | 1.30 |
|  | Males | 43.12 | 42.92 | 43.56 |  |
| Ethnic group | Bangladeshi | 0.21 | 0.24 | 0.16 | 12.03 |
|  | Black African | 0.63 | 0.63 | 0.61 |  |
|  | Black Caribbean | 0.94 | 0.96 | 0.89 |  |
|  | Chinese | 0.12 | 0.13 | 0.12 |  |
|  | Indian | 1.80 | 1.88 | 1.63 |  |
|  | Mixed | 0.85 | 0.85 | 0.85 |  |
|  | Other | 1.18 | 1.23 | 1.08 |  |
|  | Pakistani | 0.91 | 0.93 | 0.84 |  |
|  | White British | 90.31 | 90.07 | 90.85 |  |
|  | White Other | 3.04 | 3.08 | 2.97 |  |
| Socio-economic status | Higher managerial, administrative and professional occupations | 8.78 | 9.00 | 8.27 | 5.10 |
|  | Intermediate occupations | 14.44 | 14.65 | 13.96 |  |
|  | Lower managerial, administrative and professional occupations | 23.18 | 23.43 | 22.63 |  |
|  | Lower supervisory and technical occupations | 8.17 | 8.13 | 8.27 |  |
|  | Never worked and long-term unemployed | 3.62 | 3.51 | 3.86 |  |
|  | No code required | 0.12 | 0.12 | 0.13 |  |
|  | Not classified | 1.17 | 1.19 | 1.12 |  |
|  | Routine occupations | 11.84 | 11.59 | 12.40 |  |
|  | Semi-routine occupations | 16.62 | 16.44 | 17.02 |  |
|  | Small employers and own account workers | 12.06 | 11.94 | 12.34 |  |
| Disability | Limited a little | 11.48 | 11.37 | 11.73 | 5.12 |
|  | Limited a lot | 7.17 | 6.99 | 7.58 |  |
|  | Not limited | 81.35 | 81.64 | 80.69 |  |
| Highest qualification | Apprenticeship | 4.46 | 4.45 | 4.50 | 4.63 |
|  | Level 1 qualification | 18.90 | 18.62 | 19.52 |  |
|  | Level 2 qualification | 17.74 | 17.69 | 17.87 |  |
|  | Level 3 qualification | 12.77 | 12.75 | 12.80 |  |
|  | Level 4 qualification or above | 25.88 | 26.29 | 24.96 |  |
|  | No code required | 0.12 | 0.12 | 0.13 |  |
|  | No qualifications | 15.74 | 15.69 | 15.85 |  |
|  | Other qualification | 4.39 | 4.39 | 4.37 |  |
| Rural / Urban residence | Rural | 20.44 | 19.97 | 21.49 | 3.75 |
|  | Urban | 79.56 | 80.03 | 78.51 |  |
| Self-reported health | Bad | 4.66 | 4.53 | 4.94 | 3.55 |
|  | Fair | 15.80 | 15.60 | 16.24 |  |
|  | Good | 45.27 | 45.27 | 45.26 |  |
|  | Very bad | 0.99 | 0.99 | 0.99 |  |
|  | Very good | 33.28 | 33.60 | 32.57 |  |
| Index of Multiple Deprivation (IMD) Quintile Group | 1 (most deprived) | 17.82 | 17.78 | 17.92 | 5.37 |
|  | 2 | 19.31 | 19.16 | 19.65 |  |
|  | 3 | 21.08 | 20.75 | 21.82 |  |
|  | 4 | 21.63 | 21.74 | 21.37 |  |
|  | 5 (least deprived) | 19.92 | 20.35 | 18.95 |  |

**Supplementary Table 5. Distributions of covariates by time waited for treatment from decision-to-admit to admission (<18 weeks versus ≥ 18 weeks) for Other MSK conditions**

| **Characteristic** | **Level** | **All (%)** | **< 18 weeks (%)** | **≥ 18 weeks (%)** | **Absolute standardised difference (%)** |
| --- | --- | --- | --- | --- | --- |
| Age | Mean age (years) | 50.17  (SD 9.93) | 50.10  (SD 9.89) | 50.46 (SD 10.04) | 3.65 |
| Sex | Females | 54.33 | 54.83 | 52.45 | 4.78 |
|  | Males | 45.67 | 45.17 | 47.55 |  |
| Ethnic group | Bangladeshi | 0.63 | 0.65 | 0.57 | 5.21 |
|  | Black African | 1.06 | 1.05 | 1.10 |  |
|  | Black Caribbean | 1.14 | 1.13 | 1.14 |  |
|  | Chinese | 0.24 | 0.25 | 0.24 |  |
|  | Indian | 2.51 | 2.55 | 2.33 |  |
|  | Mixed | 1.32 | 1.29 | 1.41 |  |
|  | Other | 2.19 | 2.21 | 2.12 |  |
|  | Pakistani | 1.63 | 1.67 | 1.46 |  |
|  | White British | 85.25 | 85.15 | 85.63 |  |
|  | White Other | 4.04 | 4.04 | 4.01 |  |
| Socio-economic status | Higher managerial, administrative and professional occupations | 9.05 | 9.17 | 8.61 | 8.26 |
|  | Intermediate occupations | 14.39 | 14.52 | 13.92 |  |
|  | Lower managerial, administrative and professional occupations | 22.55 | 22.80 | 21.60 |  |
|  | Lower supervisory and technical occupations | 8.26 | 8.16 | 8.65 |  |
|  | Never worked and long-term unemployed | 4.50 | 4.44 | 4.72 |  |
|  | No code required | 0.61 | 0.50 | 1.00 |  |
|  | Not classified | 2.99 | 2.87 | 3.47 |  |
|  | Routine occupations | 11.27 | 11.22 | 11.47 |  |
|  | Semi-routine occupations | 15.77 | 15.77 | 15.75 |  |
|  | Small employers and own account workers | 10.60 | 10.55 | 10.81 |  |
| Disability | Limited a little | 9.08 | 9.04 | 9.25 | 4.48 |
|  | Limited a lot | 6.02 | 5.92 | 6.41 |  |
|  | Not limited | 84.90 | 85.05 | 84.34 |  |
| Highest qualification | Apprenticeship | 3.62 | 3.59 | 3.74 | 7.77 |
|  | Level 1 qualification | 18.33 | 18.22 | 18.75 |  |
|  | Level 2 qualification | 18.50 | 18.46 | 18.67 |  |
|  | Level 3 qualification | 13.95 | 13.87 | 14.24 |  |
|  | Level 4 qualification or above | 26.50 | 26.85 | 25.17 |  |
|  | No code required | 0.61 | 0.50 | 1.00 |  |
|  | No qualifications | 13.78 | 13.79 | 13.73 |  |
|  | Other qualification | 4.71 | 4.72 | 4.69 |  |
| Rural / Urban residence | Rural | 17.50 | 17.27 | 18.37 | 2.88 |
|  | Urban | 82.50 | 82.73 | 81.63 |  |
| Self-reported health | Bad | 4.21 | 4.14 | 4.48 | 2.08 |
|  | Fair | 13.60 | 13.56 | 13.74 |  |
|  | Good | 42.41 | 42.41 | 42.41 |  |
|  | Very bad | 0.94 | 0.93 | 1.01 |  |
|  | Very good | 38.83 | 38.96 | 38.35 |  |
| Index of Multiple Deprivation (IMD) Quintile Group | 1 (most deprived) | 19.31 | 19.36 | 19.11 | 5.52 |
|  | 2 | 19.89 | 19.72 | 20.56 |  |
|  | 3 | 20.50 | 20.28 | 21.30 |  |
|  | 4 | 20.55 | 20.64 | 20.23 |  |
|  | 5 (least deprived) | 19.52 | 19.79 | 18.48 |  |

**Supplementary Table 6. Distributions of covariates by time waited for treatment from decision-to-admit to admission (<18 weeks versus ≥ 18 weeks) for Other joint disorders**

| **Characteristic** | **Level** | **All (%)** | **< 18 weeks (%)** | **≥ 18 weeks (%)** | **Absolute standardised difference (%)** |
| --- | --- | --- | --- | --- | --- |
| Age | Mean age (years) | 46.75  (SD 11.07) | 47.07  (SD 10.94) | 45.69 (SD 11.43) | 12.30 |
| Sex | Females | 50.70 | 50.32 | 51.95 | 3.27 |
|  | Males | 49.30 | 49.68 | 48.05 |  |
| Ethnic group | Bangladeshi | 0.47 | 0.46 | 0.51 | 8.61 |
|  | Black African | 0.98 | 0.93 | 1.15 |  |
|  | Black Caribbean | 1.26 | 1.24 | 1.32 |  |
|  | Chinese | 0.22 | 0.22 | 0.21 |  |
|  | Indian | 2.27 | 2.30 | 2.18 |  |
|  | Mixed | 1.61 | 1.55 | 1.80 |  |
|  | Other | 1.97 | 1.96 | 2.00 |  |
|  | Pakistani | 1.28 | 1.28 | 1.29 |  |
|  | White British | 85.28 | 85.46 | 84.66 |  |
|  | White Other | 4.66 | 4.60 | 4.88 |  |
| Socio-economic status | Higher managerial, administrative and professional occupations | 9.18 | 9.47 | 8.19 | 19.46 |
|  | Intermediate occupations | 14.19 | 14.27 | 13.94 |  |
|  | Lower managerial, administrative and professional occupations | 23.04 | 23.47 | 21.62 |  |
|  | Lower supervisory and technical occupations | 8.10 | 8.23 | 7.64 |  |
|  | Never worked and long-term unemployed | 3.85 | 3.71 | 4.34 |  |
|  | No code required | 1.67 | 1.20 | 3.22 |  |
|  | Not classified | 5.30 | 4.71 | 7.25 |  |
|  | Routine occupations | 10.09 | 10.08 | 10.14 |  |
|  | Semi-routine occupations | 14.78 | 14.78 | 14.81 |  |
|  | Small employers and own account workers | 9.80 | 10.08 | 8.84 |  |
| Disability | Limited a little | 6.96 | 6.92 | 7.09 | 4.39 |
|  | Limited a lot | 4.25 | 4.16 | 4.58 |  |
|  | Not limited | 88.78 | 88.92 | 88.33 |  |
| Highest qualification | Apprenticeship | 3.45 | 3.53 | 3.17 | 15.20 |
|  | Level 1 qualification | 17.43 | 17.42 | 17.46 |  |
|  | Level 2 qualification | 18.99 | 18.83 | 19.53 |  |
|  | Level 3 qualification | 15.17 | 15.13 | 15.28 |  |
|  | Level 4 qualification or above | 28.22 | 28.68 | 26.69 |  |
|  | No code required | 1.67 | 1.20 | 3.22 |  |
|  | No qualifications | 11.01 | 11.09 | 10.72 |  |
|  | Other qualification | 4.07 | 4.12 | 3.92 |  |
| Rural / Urban residence | Rural | 17.86 | 17.75 | 18.20 | 1.18 |
|  | Urban | 82.14 | 82.25 | 81.80 |  |
| Self-reported health | Bad | 2.73 | 2.69 | 2.87 | 11.35 |
|  | Fair | 10.00 | 10.03 | 9.93 |  |
|  | Good | 39.34 | 39.80 | 37.80 |  |
|  | Very bad | 0.62 | 0.60 | 0.70 |  |
|  | Very good | 47.30 | 46.88 | 48.70 |  |
| Index of Multiple Deprivation (IMD) Quintile Group | 1 (most deprived) | 17.84 | 17.64 | 18.51 | 3.88 |
|  | 2 | 19.42 | 19.33 | 19.73 |  |
|  | 3 | 20.68 | 20.62 | 20.86 |  |
|  | 4 | 21.33 | 21.42 | 21.04 |  |
|  | 5 (least deprived) | 20.39 | 20.68 | 19.41 |  |

| **MSK condition group** | **Outcome** | **Coefficient on provider-level waiting time** | **Standard error** | **Mean individual waiting time (days)** | **Observations (n)** | **First-stage F-statistic** |
| --- | --- | --- | --- | --- | --- | --- |
| Back, neck and MSK pain | Gross pay | 0.929 | 0.015 | 83 | 190,931 | 3,992 |
|  | Employment | 0.939 | 0.013 | 83 | 191,644 | 5,353 |
|  | Benefit receipt | 0.927 | 0.025 | 80 | 42,208 | 1,358 |
| Osteoarthritis | Gross pay | 0.894 | 0.022 | 107 | 185,309 | 1,630 |
|  | Employment | 0.911 | 0.020 | 109 | 178,208 | 2,023 |
|  | Benefit receipt | 0.854 | 0.035 | 95 | 33,055 | 606 |
| Other MSK conditions | Gross pay | 0.910 | 0.025 | 86 | 208,535 | 1,349 |
|  | Employment | 0.930 | 0.017 | 87 | 207,571 | 3,096 |
|  | Benefit receipt | 0.883 | 0.028 | 79 | 31,128 | 992 |
| Other joint disorders | Gross pay | 0.906 | 0.022 | 91 | 266,605 | 1,743 |
|  | Employment | 0.924 | 0.018 | 93 | 268,183 | 2,504 |
|  | Benefit receipt | 0.912 | 0.035 | 83 | 31,589 | 693 |

**Supplementary Table 7. First-stage relationship between provider-level waiting time and individual waiting time, for the second year of modelling**

**Notes:** The dependent variable is individual waiting time from decision-to-admit to admission. The excluded instrument is the leave-one-out mean waiting time for patients treated by the same provider, in the same MSK condition group, and in the same calendar year and month of decision to admit. All specifications include the full set of covariates, baseline labour market outcomes, provider fixed effects, calendar-time fixed effects and three-digit OPCS procedure fixed effects used in the second-stage models. Standard errors are clustered by provider and year-month of decision to admit. The first-stage F-statistic tests the excluded instrument. Coefficients and standard errors are expressed on a ‘per day’ basis.

| **MSK condition group** | **Outcome** | **Coefficient on provider-level waiting time** | **Standard error** | **Mean individual waiting time (days)** | **Observations (n)** | **First-stage F-statistic** |
| --- | --- | --- | --- | --- | --- | --- |
| Back, neck and MSK pain | Gross pay | 0.887 | 0.030 | 79 | 166,354 | 857 |
|  | Employment | 0.905 | 0.029 | 80 | 166,906 | 1,004 |
|  | Benefit receipt | 0.899 | 0.030 | 79 | 33,970 | 891 |
| Osteoarthritis | Gross pay | 0.848 | 0.029 | 100 | 149,867 | 840 |
|  | Employment | 0.860 | 0.032 | 102 | 143,419 | 708 |
|  | Benefit receipt | 0.820 | 0.033 | 92 | 26,651 | 605 |
| Other MSK conditions | Gross pay | 0.880 | 0.024 | 82 | 183,011 | 1,339 |
|  | Employment | 0.895 | 0.023 | 83 | 181,590 | 1,558 |
|  | Benefit receipt | 0.834 | 0.038 | 77 | 24,885 | 474 |
| Other joint disorders | Gross pay | 0.853 | 0.028 | 87 | 237,564 | 921 |
|  | Employment | 0.869 | 0.028 | 88 | 238,191 | 994 |
|  | Benefit receipt | 0.876 | 0.032 | 82 | 25,511 | 769 |

**Supplementary Table 8. First-stage relationship between provider-level waiting time and individual waiting time, for the third year of modelling**

**Notes:** The dependent variable is individual waiting time from decision-to-admit to admission. The excluded instrument is the leave-one-out mean waiting time for patients treated by the same provider, in the same MSK condition group, and in the same calendar year and month of decision to admit. All specifications include the full set of covariates, baseline labour market outcomes, provider fixed effects, calendar-time fixed effects and three-digit OPCS procedure fixed effects used in the second-stage models. Standard errors are clustered by provider and year-month of decision to admit. The first-stage F-statistic tests the excluded instrument. Coefficients and standard errors are expressed on a ‘per day’ basis.

| **MSK condition group** | **Outcome** | **Coefficient on provider-level waiting time** | **Standard error** | **Mean individual waiting time (days)** | **Observations (n)** | **First-stage F-statistic** |
| --- | --- | --- | --- | --- | --- | --- |
| Back, neck and MSK pain | Gross pay | 0.823 | 0.041 | 75 | 145,349 | 406 |
|  | Employment | 0.823 | 0.038 | 76 | 144,088 | 458 |
|  | Benefit receipt | 0.847 | 0.040 | 78 | 25,111 | 439 |
| Osteoarthritis | Gross pay | 0.763 | 0.046 | 92 | 122,759 | 273 |
|  | Employment | 0.780 | 0.033 | 93 | 115,138 | 551 |
|  | Benefit receipt | 0.750 | 0.057 | 90 | 20,365 | 171 |
| Other MSK conditions | Gross pay | 0.819 | 0.035 | 78 | 160,743 | 555 |
|  | Employment | 0.828 | 0.029 | 79 | 157,953 | 791 |
|  | Benefit receipt | 0.777 | 0.047 | 76 | 18,573 | 275 |
| Other joint disorders | Gross pay | 0.777 | 0.048 | 82 | 212,016 | 267 |
|  | Employment | 0.791 | 0.031 | 83 | 210,258 | 634 |
|  | Benefit receipt | 0.820 | 0.056 | 80 | 19,843 | 216 |

**Supplementary Table 9. First-stage relationship between provider-level waiting time and individual waiting time, for the fourth year of modelling**

**Notes:** The dependent variable is individual waiting time from decision-to-admit to admission. The excluded instrument is the leave-one-out mean waiting time for patients treated by the same provider, in the same MSK condition group, and in the same calendar year and month of decision to admit. All specifications include the full set of covariates, baseline labour market outcomes, provider fixed effects, calendar-time fixed effects and three-digit OPCS procedure fixed effects used in the second-stage models. Standard errors are clustered by provider and year-month of decision to admit. The first-stage F-statistic tests the excluded instrument. Coefficients and standard errors are expressed on a ‘per day’ basis.

| **MSK condition group** | **Outcome** | **Coefficient on provider-level waiting time** | **Standard error** | **Mean individual waiting time (days)** | **Observations (n)** | **First-stage F-statistic** |
| --- | --- | --- | --- | --- | --- | --- |
| Back, neck and MSK pain | Gross pay | 0.902 | 0.024 | 72 | 128,890 | 1,423 |
|  | Employment | 0.816 | 0.058 | 72 | 130,957 | 199 |
|  | Benefit receipt | 0.746 | 0.063 | 76 | 15,586 | 140 |
| Osteoarthritis | Gross pay | 0.858 | 0.038 | 88 | 101,781 | 508 |
|  | Employment | 0.731 | 0.070 | 88 | 98,498 | 110 |
|  | Benefit receipt | 0.649 | 0.082 | 87 | 12,481 | 63 |
| Other MSK conditions | Gross pay | 0.902 | 0.023 | 75 | 142,197 | 1,588 |
|  | Employment | 0.801 | 0.057 | 76 | 143,410 | 199 |
|  | Benefit receipt | 0.732 | 0.069 | 75 | 11,531 | 111 |
| Other joint disorders | Gross pay | 0.869 | 0.034 | 78 | 189,629 | 664 |
|  | Employment | 0.746 | 0.091 | 79 | 193,338 | 67 |
|  | Benefit receipt | 0.747 | 0.086 | 77 | 12,682 | 76 |

**Supplementary Table 10. First-stage relationship between provider-level waiting time and individual waiting time, for the fifth year of modelling**

**Notes:** The dependent variable is individual waiting time from decision-to-admit to admission. The excluded instrument is the leave-one-out mean waiting time for patients treated by the same provider, in the same MSK condition group, and in the same calendar year and month of decision to admit. All specifications include the full set of covariates, baseline labour market outcomes, provider fixed effects, calendar-time fixed effects and three-digit OPCS procedure fixed effects used in the second-stage models. Standard errors are clustered by provider and year-month of decision to admit. The first-stage F-statistic tests the excluded instrument. Coefficients and standard errors are expressed on a ‘per day’ basis.

**Supplementary Figure 1. Study sample flow diagram**

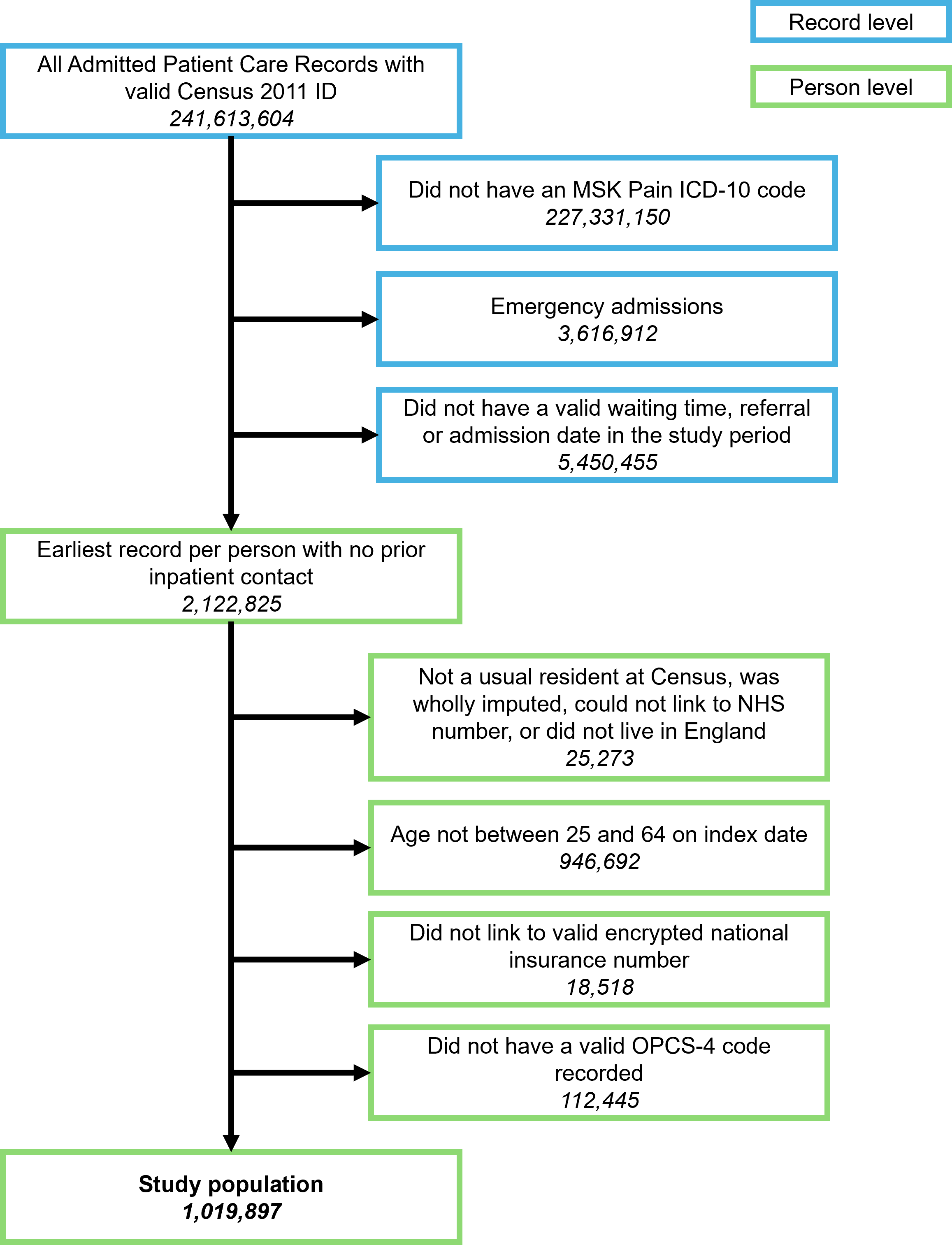

**Supplementary Figure 2. Estimated relationship between waiting time and gross employee pay up to five years after treatment, stratified by MSK condition group and sex**

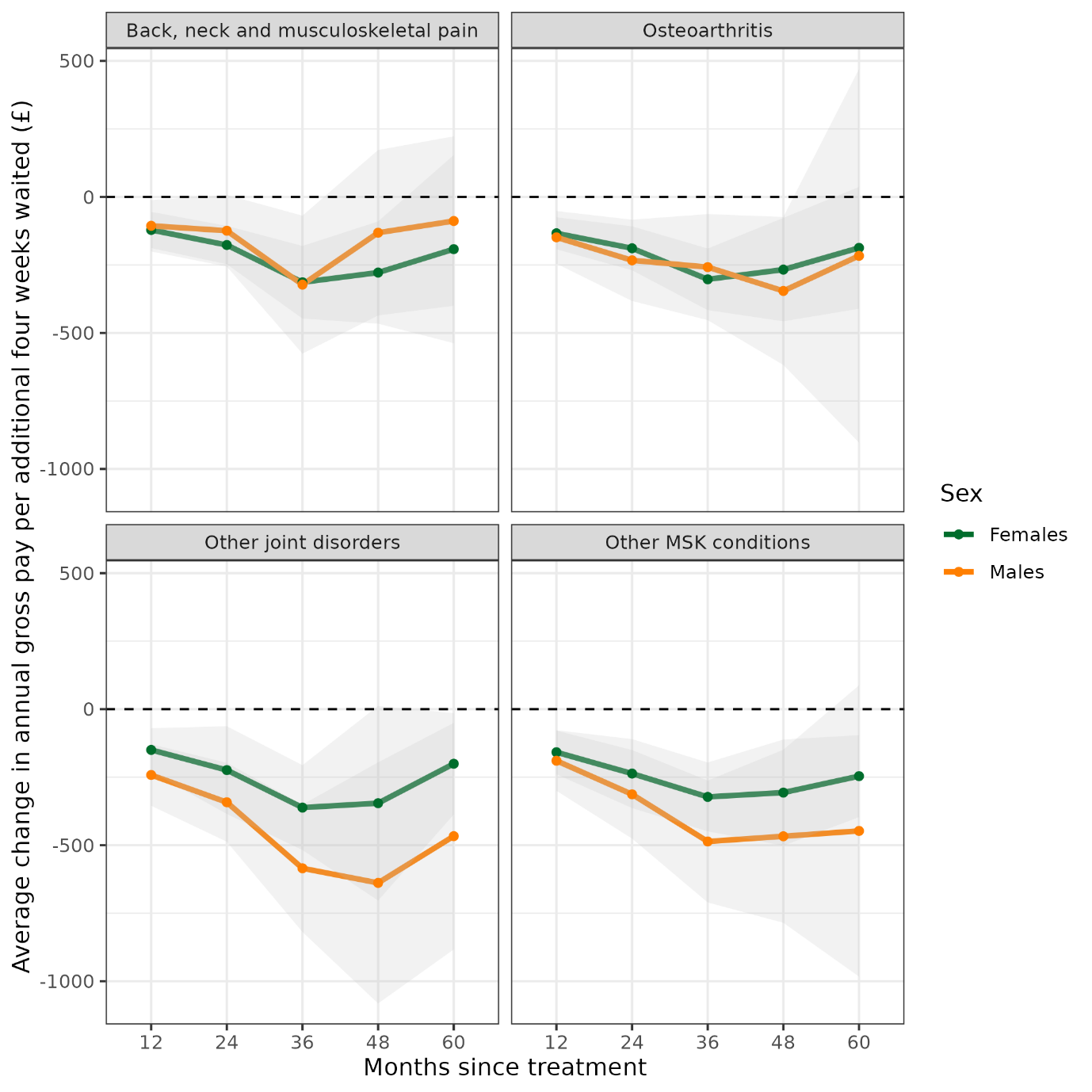

*Notes: Estimates are adjusted for age at index date, sex, ethnic group,* socio-economic status, disability, highest qualification, rural or urban residence, self-reported general health and IMD quintile group, baseline pay, and fixed effects (provider, referral year and month, and three digit OPCS-4 code)..

*Shaded areas are 95% confidence intervals. Values are expressed in 2023 prices.*

**Supplementary Figure 3. Estimated relationship between waiting time and gross employee pay up to five years after treatment, stratified by MSK condition group and age group**

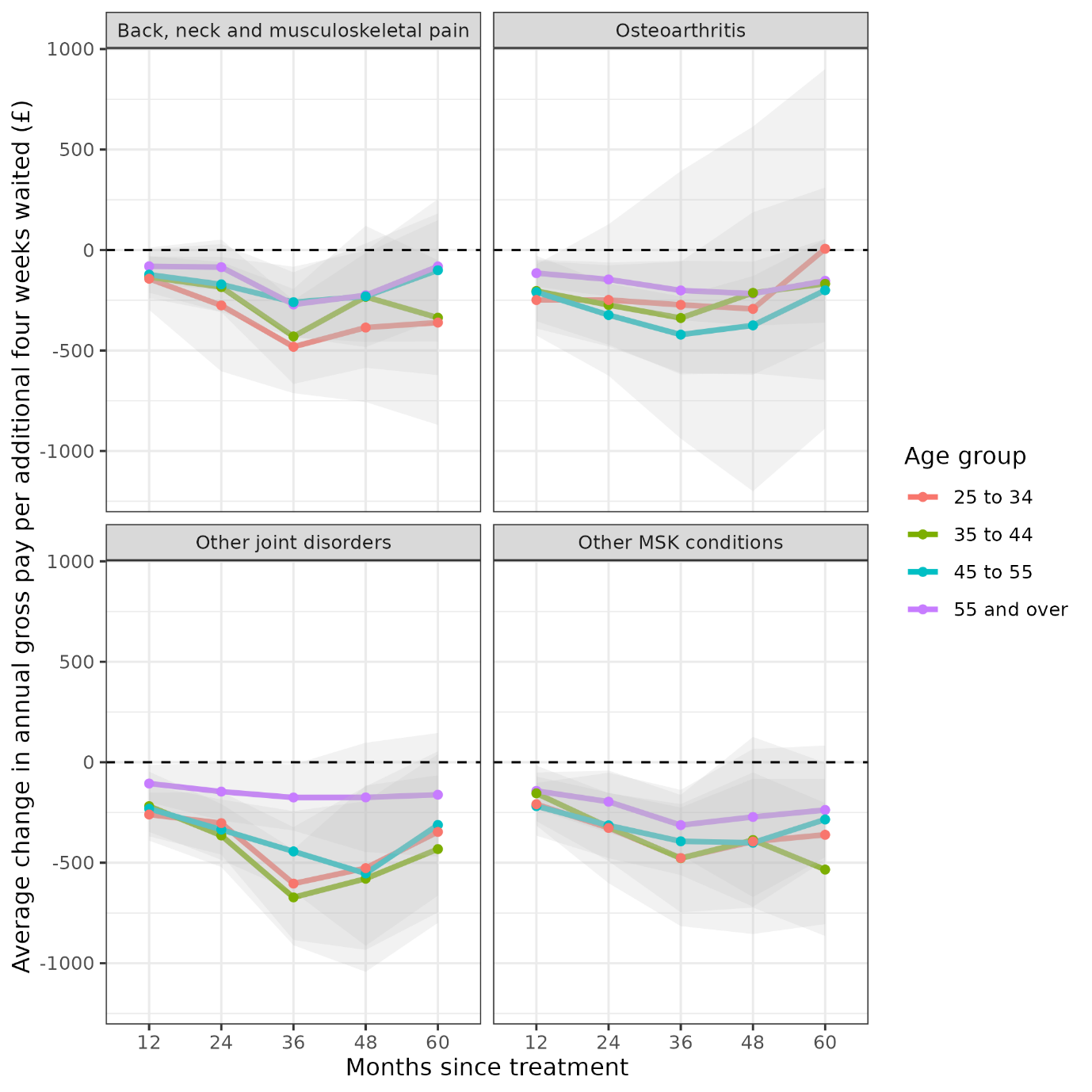

*Notes: Estimates are adjusted for age at index date, sex, ethnic group,* socio-economic status, disability, highest qualification, rural or urban residence, self-reported general health and IMD quintile group, baseline employee status, and fixed effects (provider, referral year and month, and three digit OPCS-4 code)..

*Shaded areas are 95% confidence intervals. Values are expressed in 2023 prices.*

**Supplementary Figure 4. Estimated relationship between waiting time and gross employee pay up to five years after treatment, stratified by MSK condition group and area deprivation quintile group**

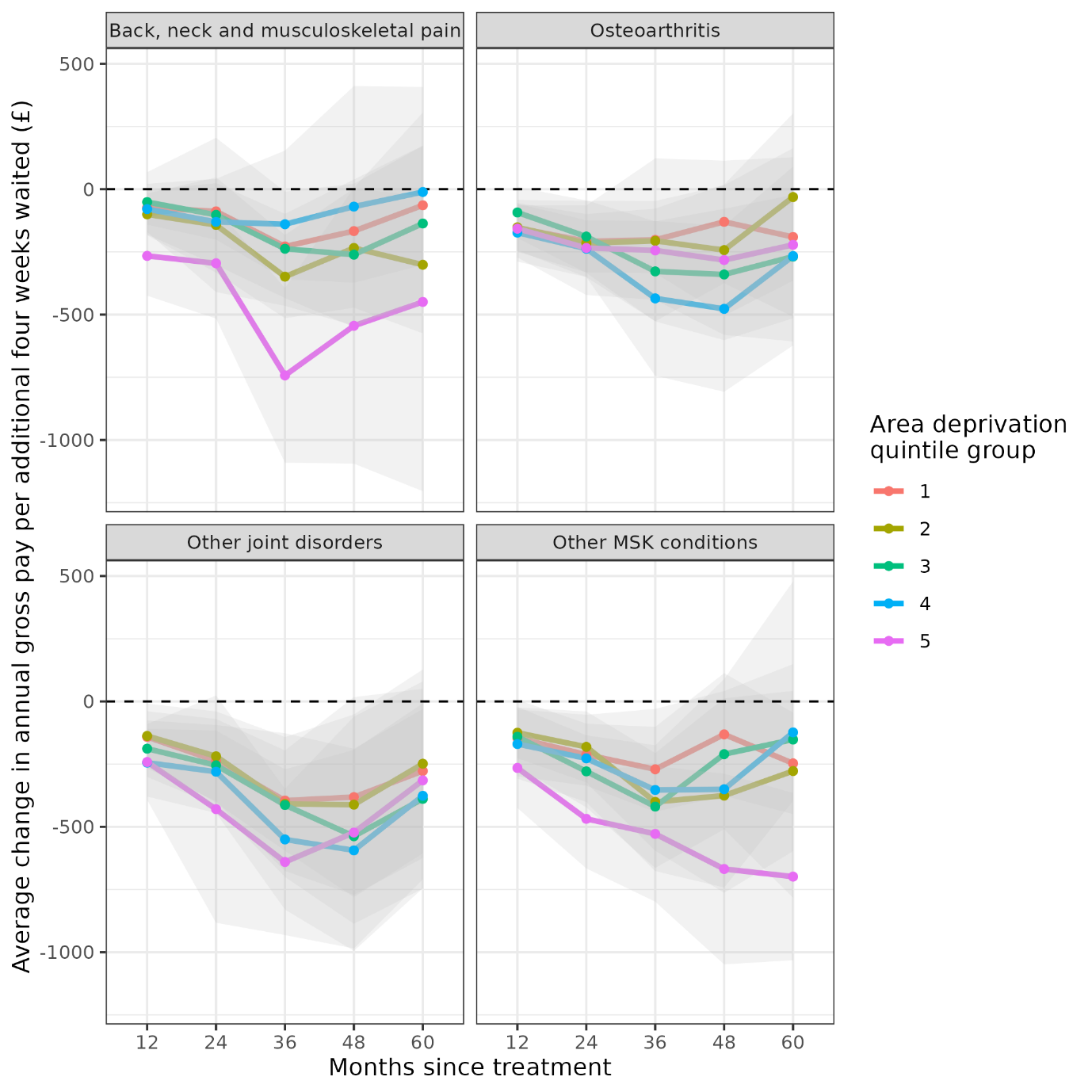

*Notes: Estimates are adjusted for age at index date, sex, ethnic group,* socio-economic status, disability, highest qualification, rural or urban residence, self-reported general health and IMD quintile group, baseline benefit receipt status, and fixed effects (provider, referral year and month, and three digit OPCS-4 code)..

*Shaded areas are 95% confidence intervals. Values are expressed in 2023 prices.*

**Supplementary figure 5. Cumulative Distribution Function (CDF) plot**

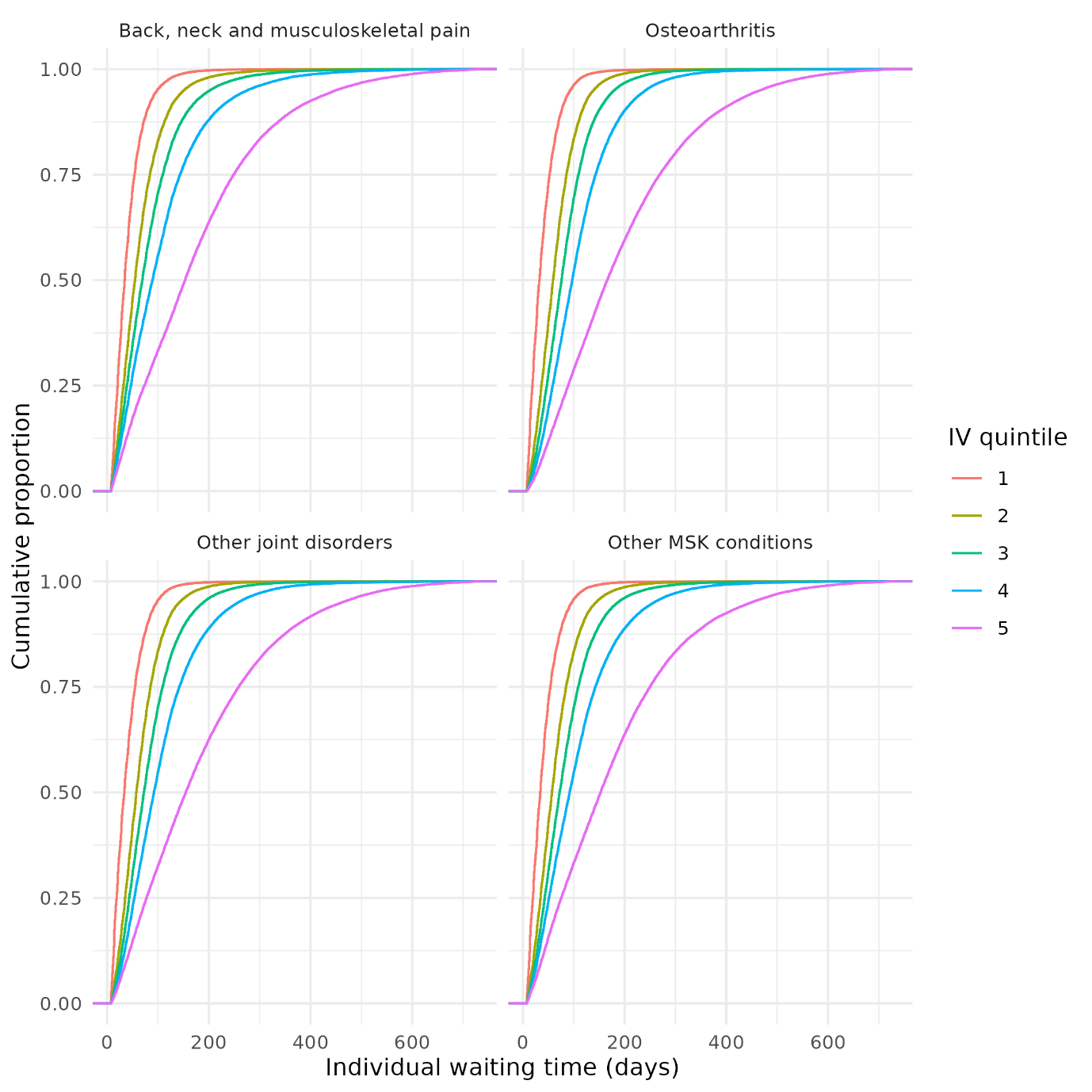

**Supplementary figure 6. Assessment of linearity in the instrument-exposure relationship, quadratic polynomial fit**

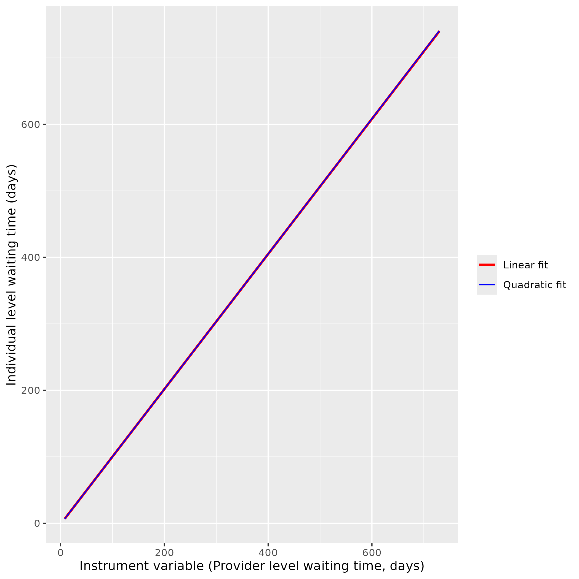

**Supplementary figure 7. Assessment of linearity in the instrument-exposure eelationship, cubic polynomial fit**

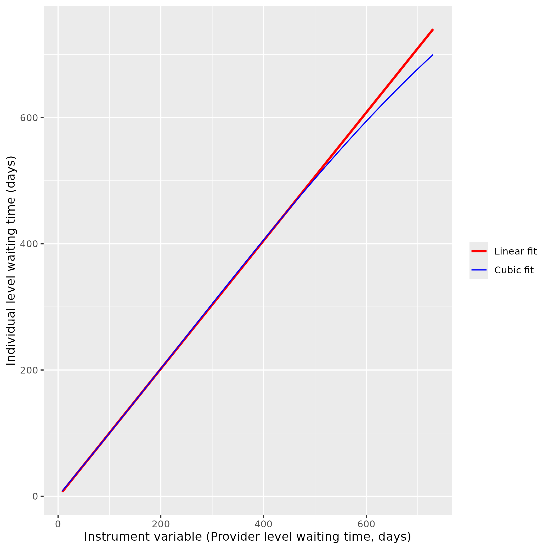

**Supplementary Figure 8. Assessment of linearity in the instrument-exposure eelationship, spline fit**

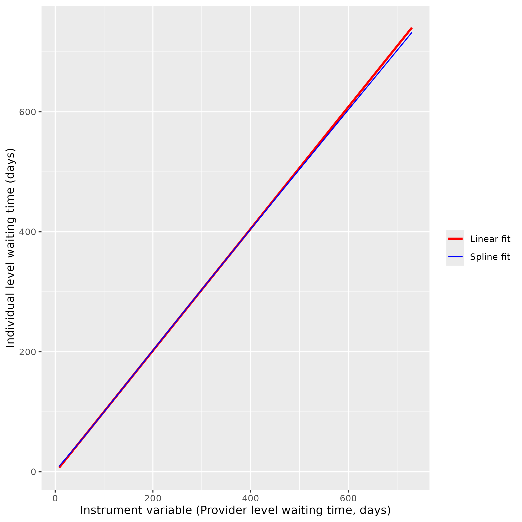

**Supplementary Figure 9. Estimated relationship between individual level waiting time (no IV) and pay up to five years after treatment, stratified by MSK condition group**

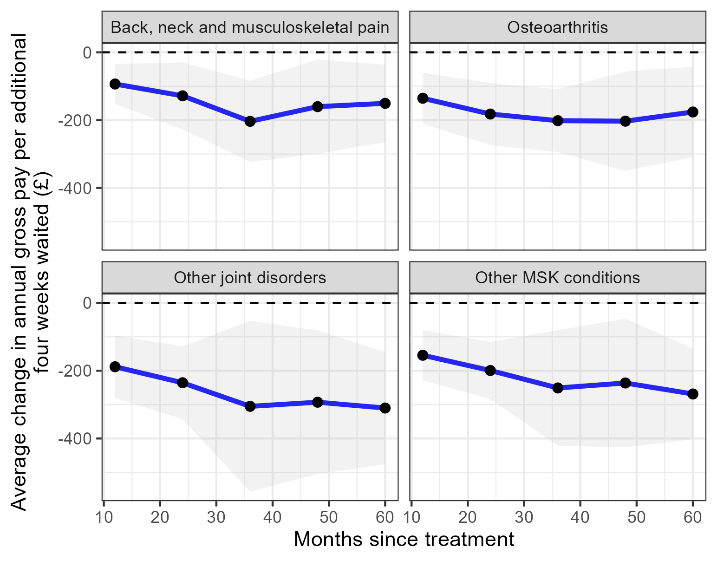

**Supplementary Figure 10. Estimated relationship between individual level waiting time (no IV) and employment status up to five years after treatment, stratified by MSK condition group**

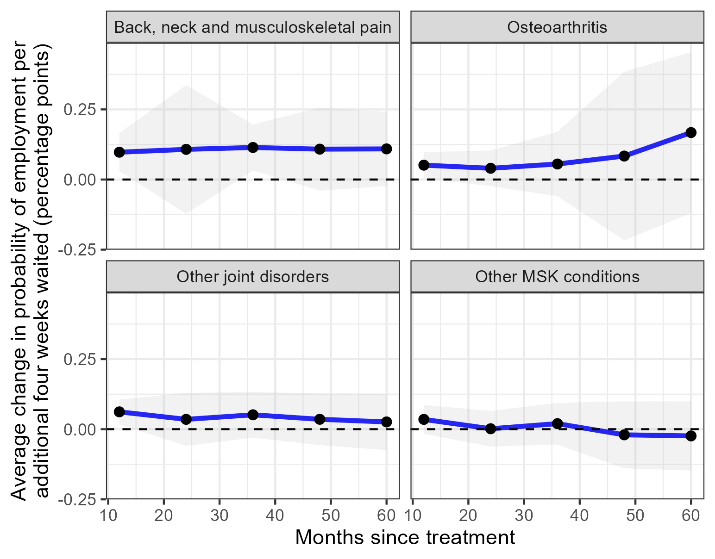

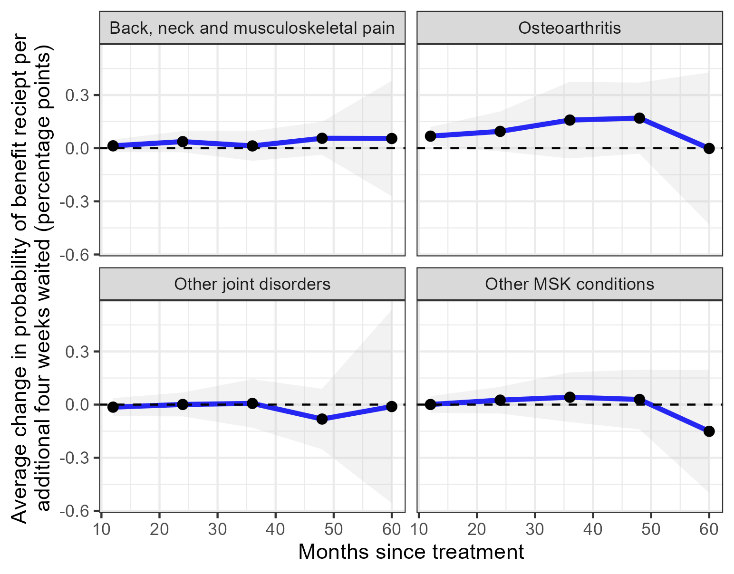
**Supplementary Figure 11. Estimated relationship between individual level waiting time (no IV) and benefit receipt up to five years after treatment, stratified by MSK condition group**

**Supplementary Figure 12. Estimated relationship between waiting time and pay up to five years after treatment, stratified by MSK condition group, with provider and procedure interaction term included as a fixed effect and in the clustered standard errors**

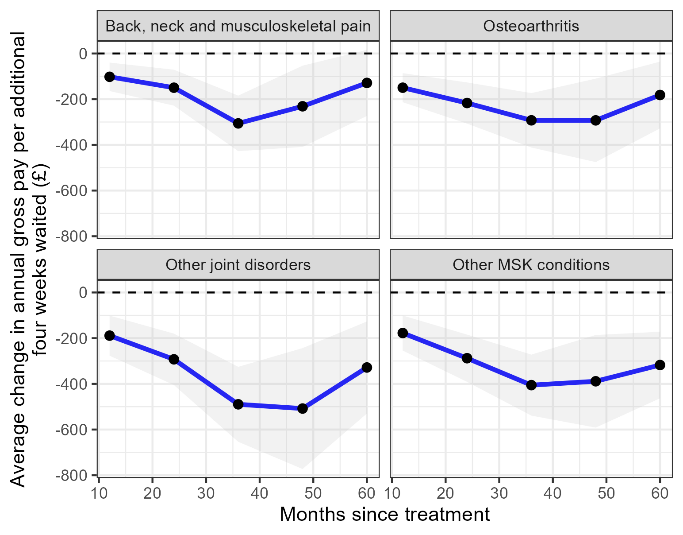

**Supplementary Figure 13. Estimated relationship between waiting time and employment status up to five years after treatment, stratified by MSK condition group, with provider and procedure interaction term included as a fixed effect and in the clustered standard errors**

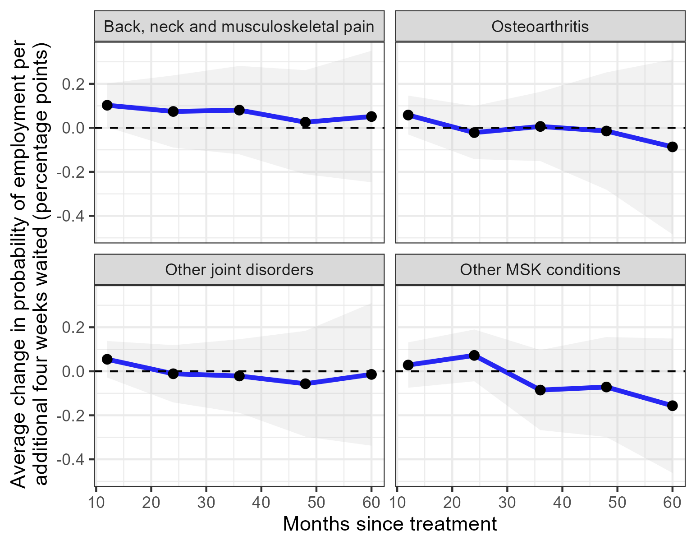

**Supplementary Figure 14. Estimated relationship between waiting time and benefit receipt up to five years after treatment, stratified by MSK condition group, with provider and procedure interaction term included as a fixed effect and in the clustered standard errors**

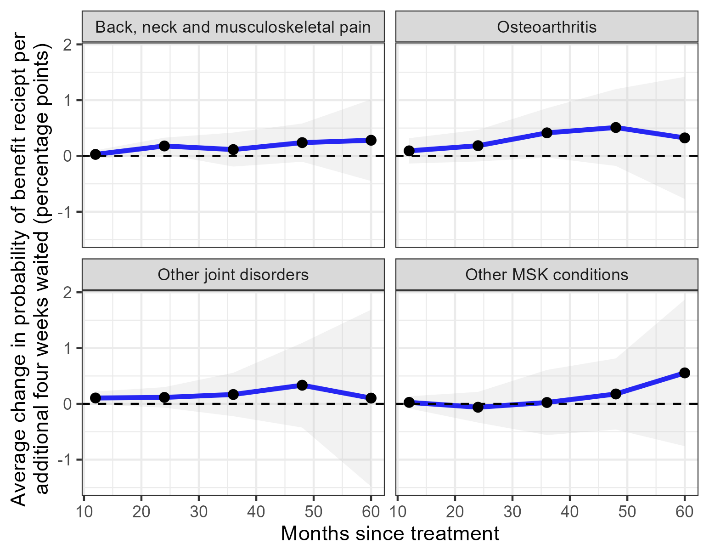

**Supplementary Figure 15. Estimated relationship between waiting time and pay up to five years after treatment, stratified by MSK condition group, without individual characteristics as covariates**

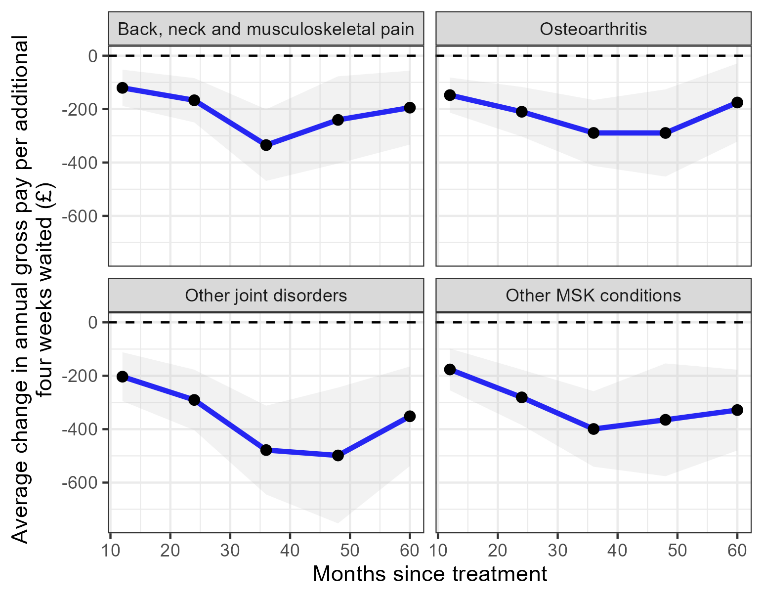

**Supplementary Figure 16. Estimated relationship between waiting time and employment status up to five years after treatment, stratified by MSK condition group, without individual characteristics as covariates**

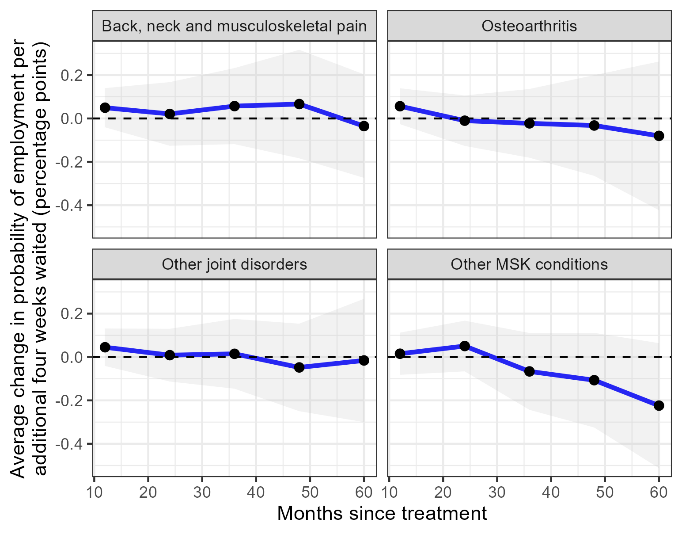

**Supplementary Figure 17. Estimated relationship between waiting time and benefit receipt up to five years after treatment, stratified by MSK condition group, without individual characteristics as covariates**

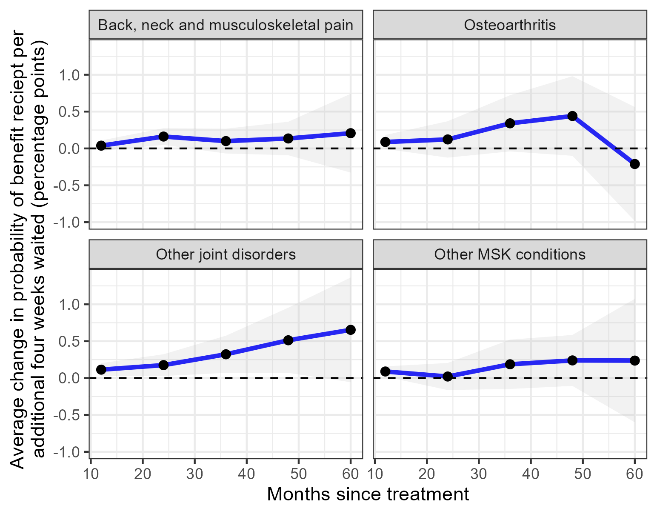

**Supplementary Figure 18. Estimated relationship between waiting time and pay up to five years after treatment, all MSK condition groups combined**

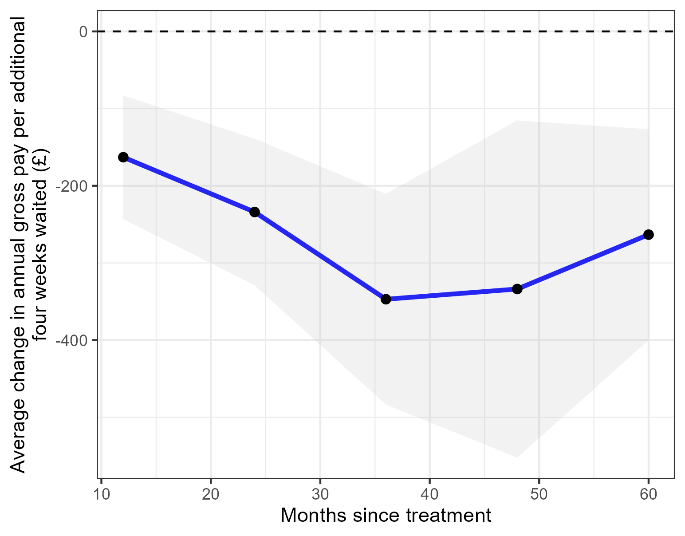

**Supplementary Figure 19. Estimated relationship between waiting time and employment status up to five years after treatment, all MSK condition groups combined**

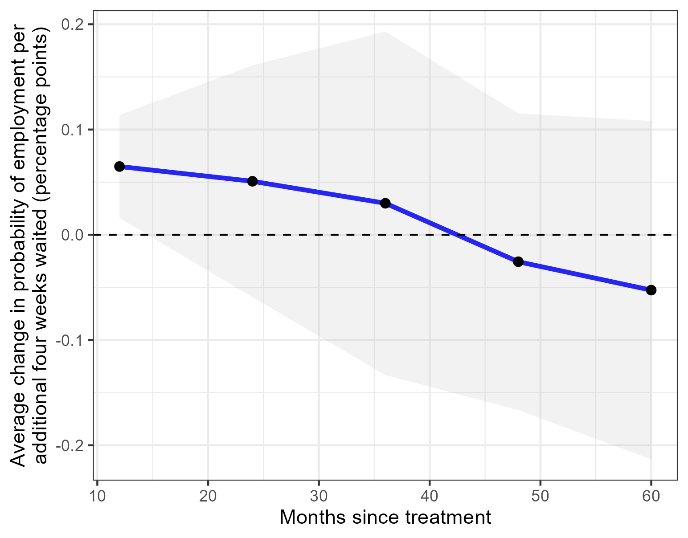

**Supplementary Figure 20. Estimated relationship between waiting time and benefit receipt up to five years after treatment, all MSK condition groups combined**

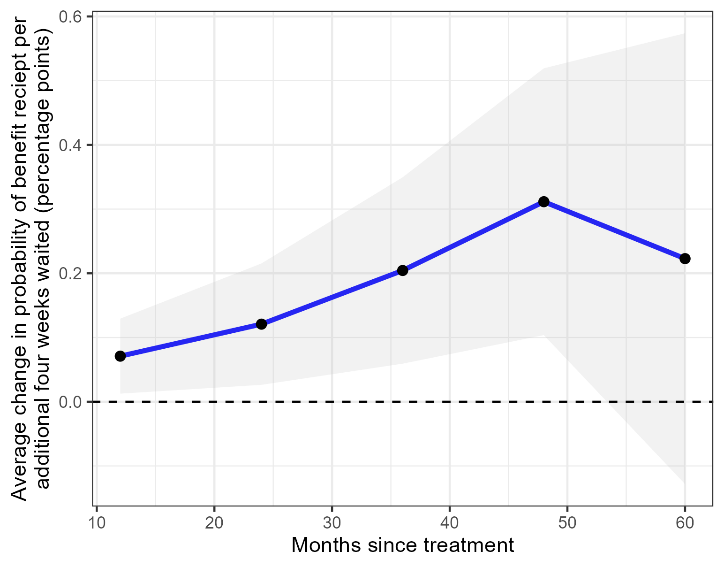
